# Random effects adjustment in machine learning models for cardiac surgery risk prediction: a benchmarking study

**DOI:** 10.1101/2023.06.08.23291129

**Authors:** Tim Dong, Shubhra Sinha, Daniel P Fudulu, Jeremy Chan, Ben Zhai, Pradeep Narayan Narayan, Massimo Caputo, Andy Judge, Arnaldo Dimagli, Umberto Benedetto, Gianni D. Angelini

**Affiliations:** Bristol Heart Institute, Translational Health Sciences, University of Bristol; Department of Cardiac Surgery, Rabindranath Tagore International Institute of Cardiac Sciences, India; Department of Computing Science, Northumbria University

**Keywords:** cardiac surgery, artificial intelligence, risk prediction, machine learning, operative mortality, confounding analysis, causal analysis, performance analysis, national dataset

## Abstract

**Objectives:** There is an ongoing debate over whether a procedural specific (e.g. Society of Thoracic Surgeons (STS)) or universal model (e.g. EuroSCORE II (ES II)) should be used for patient selection in cardiac surgery. Recently, we showed that ES II suffers from severe performance drift across several important metrics and that ML approaches such as Xgboost and Random Forest are substantially more resistant to dataset drift. With the growing interest in big data and its leverage through the use of ML approaches that are not limited by linear statistical assumptions, the number of clinical variables can theoretically increase exponentially. In addition, the variations and residual confounding that historically hindered the usefulness of cardiac risk stratification scores can potentially be taken into account. Here, we assess these possibilities on a large United Kingdom (UK) database.

**Methods:** A retrospective analysis of prospectively routinely gathered data on adult patients undergoing cardiac surgery in the UK between 2012-2019. We temporally split the data 70:30 into a training and validation subset. Two sets of seven ML mortality prediction models, with and without variable selection were assessed for consensus Clinical Effective Metric (CEM) overall performance and performance within each of CEM’s consistuent metrics. Confounding and potential causal relationships between covariates and outcomes were evaluated using bayesian network analysis.

**Results:** A total of 227,087 adults underwent cardiac surgery during the study period with a mortality rate of 2.76%. For non-variable selected (NVS) risk scores with 102 variables, Xgboost with adjustment for hospital variation was superior to the Xgboost without adjustment (p < 2e-16). Both NVS and the 18 variables selected (VS) Xgboost with adjustment for hospital variation risk scores were superior to the Xgboost (ES II 18 variables) model (p < 6.3e-15), with NVS Xgboost with adjustment for hospital variation having the best performance, followed by the VS Xgboost with adjustment for hospital variation (CEM Difference: 0.0150 and 0.0023, respectively).

**Conclusions:** We have identified an ML adjusted risk score comprising 102 variables that increases risk stratification performance on hold out dataset, removing the need to perform variable selection and reduction. This paves the way for further research that utilises this new set of variables with hospital-based adjustments for the safer selection of patients undergoing cardiac surgery.

## Introduction

The importance of Machine Learning (ML), a branch of Artificial intelligence (AI) has recently been highlighted as a potential alternative to mortality risk scores for cardiac surgical procedures, such as Society of Thoracic Surgeons (STS),[1] and EuroSCORE II (ES II) [2] which are prone to miscalibration overtime and poor generalisation across datasets.[1,3] In particular, ES II, which is based on logistic regression using 18 items of patient information, has been shown by numerous studies to display poor discrimination and calibration across datasets with differing characteristics, including but not limited to age,[4] ethnicity[5] and procedures groups.[6–10] Furthermore, ES II suffers from severe performance drift across several metrics including but not limited to discrimination, calibration, clinical utility and overall accuracy. ML approaches such as Xgboost and Random Forest are substantially more resistant to performance drifts that arise as a consequence of dataset drift.

ML has been shown to be superior to conventional scoring systems with the magnitude and clinical influence of such improvements demonstrated.[2] The ability to counter-performance drift due to temporal changes in the prevalence of risk factors has also been evaluated across multiple centres and has been shown to be superior to universal scores such as the ES II. However, the confounding effects of variables not included in ES II for its consistuent procedures have not been taken into account and may confuse correlation with causation of the outcome,[11] and may also limit the full potential of the risk stratification scores. The influence of these “hidden” variables on the performances of ML scores have yet to be fully elucidated.

Parsimonious models can result in improved prediction by preventing overfitting in scenarios where the number of events to variables are low, e.g. small sample size and high dimensional datasets. However, The No Free Lunch Theorem states that all optimisation algorithms perform equally well when their performance is averaged across all possible problems,[25] and suggests that different ML models will have a different set of optimal prediction variables for any given task or dataset. This makes it difficult to provide a fair comparison of models based on equal events per variable (EPV). Keeping the EPV constant would require different sample size of data being compared due to differences in number of variables per model. This would lead to unfairness due to sample size disparity. An alternative solution to create a comparative set of parsimonious models is to keep EPV constant by using the top *N* most important variables across all models, thereby also fixing the sample size across all model comparisons.

Since cardiac surgery mortality events are low and typically in the order of 3%, the events per variable (EPV) will require careful judgement of the right balance of variables to use for prediction modelling.[12] In small datasets, the cardiac surgery risk predictions are more likely to be biased and to have high error rates, especially when larger numbers of variables are used.[13] The commonly used ES II is limited to the use of 18 variables partly for this reason. However, for larger multi-centre National Adult Cardiac Surgery Audit (NACSA) datasets, the number of events will be larger relative to smaller datasets, theoretically supporting the use of larger number of variables. One other reason that ES II does not use a larger number of variables is due to restrictions in logistic regression that variables should be absent of multi-collinearity i.e. predictors should be independent of each other. This assumption becomes more difficult to meet as variable numbers increases. However, ML is not affected by this limitation and is more suited to modelling complex non-linear relationships among variables. We therefore hypothesised whether ML could be applied to enable increased performance when a larger set of variables are used.

One other issue of using a larger multi-centre national dataset such as NACSA is that there may exist systematic differences in relationships of variables across different centres, i.e. calibration drift across geographic regions. For example, each hospital could have different cardiac surgery protocols or different suppliers for surgical equipments and devices. In addition, patients from different centres are likely to have different levels of deprivation and social demographic profiles that could result in regional differences in cardiac surgery risk. Therefore, we hypothesise that taking hospital variation into account could improve the performance of cardiac surgery risk prediction.

We, therefore, trained and evaluated two sets of 7 supervised ML models based on various combinations of a large set of 60 clinical variables with and without hospital location variable(s) to (1) determine the best ML model in terms of overall accuracy, discrimination, calibration and clinical effectiveness, (2) use variable importance to select and build a parsimonious version of the set of 7 models from (1) and to compare performances within and across parsimonious and non-parsimonious sets; (3) analyse causal and non-causal relationships between newly proposed variables and the outcome.

## Related works

Machine learning (ML) techniques have drawn interest as potential substitutes for existing scoring systems for predicting the mortality risk of cardiac surgery. ML models have been demonstrated to perform better than traditional methods, offering predictions that are more accurate with potential treatment optimization applications. However, prior studies of cardiac surgery risk prediction modelling using ML have mostly concentrated on applying ML algorithms without taking into account the potential variability of patients across various hospital locations.

Incorporating random effects into machine learning models for risk prediction applications has drawn attention in recent years. By incorporating random effects, this method, sometimes referred to as mixed effects machine learning, may take into account the heterogeneity introduced by various hospital locations. The model can capture the systematic variations in the interactions between variables across different centres through the incorporation of random effects, which enhances the risk prediction model’s overall performance and calibration.

While one survival analysis study using ML on cardiovascular related risk factors highlighted the importance of using the hospital index as a random effect to account for clustering within the hospital,[14] this random effects was only evaluated for the traditional Cox proportional-hazards regression as part of the sub-analysis without similar adjustment in the Random Forest model. While there are few studies specific to mixed machine learning in cardiac surgery, one study in the related field of cardiovascular imaging applied linear mixed effects model as a post-processing step following convolutional neural network (CNN) to compare across different coefficient of variation across decision made by human and the CNN.[15] However, the no mixed effects machine learning or deep learning model was used.

In other domains such as neurology[16] mixed effects linear regression have also been used to account for random variations across models compared. However, such studies also apply mixed effects separate to the main models analysed rather than incorporating effects with the machine learning model. Unlike previously mentioned studies which applied linear mixed effect models to compare machine learning models, another study in precision oncology used random effects within a linear mixed model to select features while adjusting for patient variations.[17] In a different study on plant sciences utilising a random forest, similar linear mixed random effects selection of features before machine learning modelling was used.[18]

We have previously evaluated the calibration changes in machine learning base models across the 1996-2011 and 2011-2017 for the EuroSCORE I variables and shown that both LR and random forest models were associated with good discrimination ability but substantial miscalibration.[19] We followed this by a development of a suit of performance metrics for evaluating clinical machine learning models.[20] In a separate study, we have developed an approach that compared calibration changes, variable importance drift, performance drift and actual dataset drift of the base models using EuroSCORE II variables across the years 2017-2019.[21] With respect to changes in techniques, Dataset drift was observed across the Holdout time periods for Weight of intervention of EuroSCORE II. Sharp dataset drifts were observed for the Single non-CABG and 3 procedures category between 2018-12 to 2019-02. In a separate study, we found that machine learning models could be ensembled to combine EuroSCORE I and EuroSCORE II variables and data from different time periods to improve performance through Xgboost homogeneous ensembles.[22]

While prior research has emphasised the need for mixed effects in machine learning studies and related techniques in prediction, little research has specifically compared the different ways of encoding random effects to account for hospital site heterogeneity in ML models. By using a mixed effects machine learning approach with random effects as hospital location, this research aims to bridge this gap. The suggested model can effectively account for regional variations in cardiac surgery operation and patient characteristics by taking into consideration the hospital-specific impacts. As a result, the model’s predictions of mortality risk are more precise and reliable.

## Materials and methods

The register-based cohort study is part of a research approved by the Health Research Authority (HRA) and Health and Care Research Wales and a waiver for patients’ consent was waived (HCRW) (IRAS ID: 278171). An Abbreviations and Definitions list of frequently used technical terms used in this study has been provided for the reader at the start of the Supplementary Materials.

### Dataset and Patient Population

The study was performed using the National Adult Cardiac Surgery Audit (NACSA) dataset, which comprises data prospectively collected by National Institute for Cardiovascular Outcome Research on all cardiac procedures performed in all NHS hospital sites and some private hospitals across the UK.[19]

Patients undergoing cardiac surgery from 42 cardiac surgery centres between 1 Jan 2012 and 31 Mar 2019 were included. Missing and erroneously inputted data in the dataset were cleaned according to the National Adult Cardiac Surgery Audit Registry Data Pre-processing recommendations;[23] details are found in the Supplementary Materials, Table S1: Handling of missing data and Supplementary Materials

Treatment of Missing Data section. Missing categorical variable values were generally set to the baseline level, i.e., no risk were present, except where other specific values are more appropriate. Missing continuous variable values were imputed using the median (Hmisc R package). Detailed variable processing are shown in Table S1. Variable distributions were checked using histogram plots. Data standardization was performed by subtracting variable mean and dividing by the standard deviation values.[24]

The dataset was split into two cohorts: Training/Validation (n = 157196; 2012-2016) and Holdout (n = 69891; 2017-2019) as per previous studies.[20,21]

### Baseline Statistical analysis

Continuous variables are compared using non-parametric Wilcoxon rank-sum test, whilst categorical variables are compared using Pearson’s χ tests or Fisher’s exact test as appropriate. Baseline variable characteristics were assessed by pooling the top 18 most important variables identified through SHAP from each model and retaining only unique variables across all models.

Scikit-learn v0.23.1 and Keras v2.4.0 were used to develop the models and to evaluate their discrimination, calibration and clinical effectiveness capabilities. Statistical analyses are conducted using STATA-MP version 17 and R v4.0.2.[25] Anova Assumptions were checked using R rstatix package.

### Variable Selection and Processing

A total of 245 NACSA variables were considered. 179 indication, intra-op, anatomical, rare procedures, dates, comorbidities, other outcomes and similar variables were excluded. An additional 3 intra-op variables were excluded resulting in 63 variables. Two variables (ethnicity and the total number of grafts) were excluded because they were only recorded at 1 hospital. Intra-Aortic Balloon Pump usage (IABP) and ventricular assist device used (VAD) were recoded to two levels (pre-op usage: Yes or No). Aortic valve (AV), Mitral Valve (MV), Tricuspid Valve (TV) and Pulmonary valve (PV) procedures were recoded to two levels (procedure performed: Yes or No). Pre-op sinus, Atrial Fibrillation (AF), Ventricular Fibrillation or Tarchicardia, and heart block or paced rhythm were combined as a single categorical variable with levels 0, 1, 2, 3, respectively. Aortic, Tricuspid, Pulmonary and Mitral valve procedures were made more general by combining individual repair types into a single repair category, resulting in three levels: 0.None; 1.Repair; 2.Replacement. Further variable processing details are provided in Table S1, resulting in 61 initial sets of variables (Figure S2). A correlation analysis was conducted to determine the collinearity of variables.

### Fixed and mixed effects dataset

Three different versions of datasets were generated based on whether the geographic location of the cardiac centre was 1) modelled as mixed-effects model or 2) not; 3) excluded. The number of baseline variables for 1) is 61; 2) is 60 + 42 = 102; and for 3) is 60.

1) Mixed Effects modelling The geographical location of the 42 centres was converted from the character format to a single vector of numeric equivalents and entered as the random effects.
2) Fixed Effects modelling The geographical location of the 42 centres was hot-encoded into 42 new variables each indicating whether or not each procedure originated from that geographical location.
3) No Cardiac Centre This is the baseline model with the removal of the geographical location variable.

### Non-variable selected Modelling

Using the above datasets, seven models were developed without variable selection. Those model included Xgboost – centres,[26] Xgboost + centres (hot encoded), RF – centres,[27] RF + centres (hot encoded), Mixed Effects RF (MERF) + random effects (RE): centre, Mixed Effects Xgboost + RE: centre and GPBoost + RE: centre, where – indicates the exclusion of centre variable and + indicates inclusion of centre variable. Due to the MERF and Mixed-effects Xgboost requiring the outcome to be in a continuous format, the outcome variable for these two models was transformed into probabilities based on the corresponding training set. Xgboost – centres was considered as the baseline comparison.

### Variable selected Modelling

Shapley global variable importance was used to identify the top 18 most important variables for each of the above models respectively.[28] *N* = 18 was chosen in so as to enable comparison to the performance of ES II and ML models built using ES II. The identified variables was used to re-build a parsimonious version of the above set of seven models. Due to the MERF and Mixed effects Xgboost requiring the outcome to be in continuous format, the outcome variable for these two models was transformed into probabilities based on the corresponding variable selected training set. Xgboost – centres was considered as the baseline comparison.

Further details on model development can be found in Supplementary Materials, section: Model Specification.

### Hyperparameter Tuning

For non-variable selected models, a shuffled and stratified randomized 3-fold cross-validation (CV) search on hyperparameters was conducted for Xgboost – centres, Xgboost + centres (hot-encoded), RF – centres, RF + centres (hot-encoded). The process was repeated for the top 18 variables identified from shapley for each corresponding model. For non-variable selected models, the optimal hyperparameters for RF – centres and Xgboost – centres were used as initial hyperparameters for the Mixed-effects RF (RE: centre) and Mixed-effects Xgboost (RE: centre), respectively and the maximum number of expectation maximization (EM) iterations was set to five. For variable selected models: Mixed-effects RF (RE: centre) and Mixed-effects Xgboost (RE: centre) models, optimal hyperparameter values were identified using RF and Xgboost with shuffled and stratified randomized 3-fold cross-validation (CV) search on the top 18 variables from the corresponding non-variable selected models. Shuffled and stratified CV hyperparameter tuning was not possible for GPBoost, so a randomised 3-fold CV search on hyperparameters was conducted. In order to determine the optimal hyperparameters from the set of possible parameters, including parameters informed by previous studies,[20] 30 different combinations were randomly selected and evaluated in each fold of CV.

This hyperparameter selection process was conducted for both the non-variable selected and variable selected sets of models.

### Assessment of model performance

External validation was performed on the Holdout dataset (2017-2019).[29] Each model calculated the probability of surgical mortality for each patient. As per previous studies,[21,22] we applied the consensus metric approach of Clinical Effectiveness Metric (CEM), using the combined geometric average results of all metrics[30]:

1. Discrimination: AUC[31], F1 score[32]
2. Calibration: 1 – ECE.[33]
3. Overall accuracy[30]: 1 – Brier score.[34]
4. Clinical utility – Net benefit Analysis[35]

Further details on the theory and application of each individual metric above is provided in our previous study of machine learning models using ES II variables.[20] One thousand bootstrap samples were taken for the CEM and its constituent metrics. Further details of this metric can also be found in the Supplementary Materials, section: Assessment of model performance.

We evaluated the following comparisons:

1) Non-variable selected Xgboost – centres model against all other non-variable selected models.
1) Variable selected Xgboost – centres model against all other variable selected models.
1) Highest performing model from 1) against that from 2) and Xgboost built using EuroSCORE II (ES II) variables, with the latter model as the control.[ref prj 1.2]

For comparisons 1) and 2), adapted Rain plots in R-3.6.2 was used to visualise constituent metrics within CEM.[36] For comparisons 1), 2) and 3), differences across models’ CEMs were tested using Repeated measures One-Way Anova and Bonferroni Corrected multiple pairwise paired t-tests; this was followed by Dunnett’s Correction for multiple comparisons. ANOVA assumptions for outliers were checked. Normality assumptions were checked using the Shapiro-Wilk test and histogram plots.[37] An overview of the study design is shown in Figure 1.

**Figure 1.**
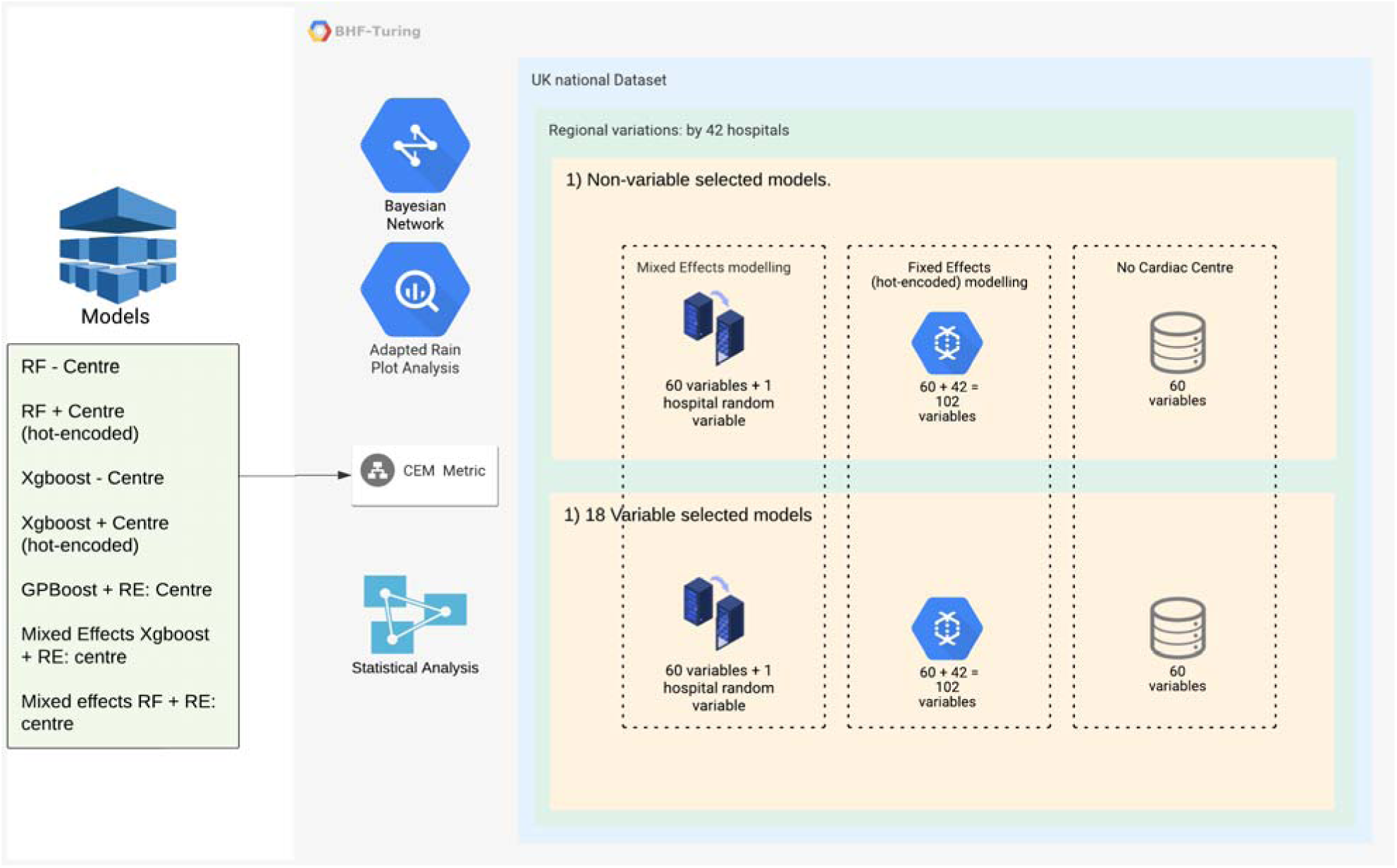
Design overview of the study; Non-variable and variable selected analyses are performed; CEM is used to simultaneously assess discrimination, calibration, clinical utility, overall accuracy; analyses are performed for i) Mixed effects modelling with 60 main variables and hospitals as a single random variable; ii) Fixed effects modelling with 60 main variables and 42 hospitals each as a single variable and iii) Without consideration of hospital variation.

### Bayesian Networks

A Bayesian network is produced using the training dataset to interpret 1) the relationship between the 18 most important variables and the outcome, for the highest overall performing model and;[38] 2) the relationships between variables in 1) and the other variables used for the non-variables selected models.

## Results

### Patients characteristics

A tota of 227,087 procedures of patients over 18 years of age from 42 UK hospitals were included in this analysis, following the removal of 3,930 congenital cases, 1,586 transplant and mechanical support device insertion cases and 3,395 procedures missing information on mortality (Table 1). There were 6,258 deaths (mortality rate of 2.76%). The primary outcome of this study was in-hospital mortality. A CONSORT flow diagram is shown in Figure S1.1. Missing rates of variables were low except for Left Ventricular Function, pulmonary artery systolic pressure (PAsys), and Number of Valves (Figure S1.2). Missing variables were backfilled using other informative variables according to NACSA dataset cleaning protocol: https://www.nicor.org.uk/wp-content/uploads/2018/09/nacsacleaning10.3.pdf and then imputed to improve variable quality, after which there were no missing variable values.

**Table 1.** Patient Demographics. Summary of cleaned and pooled top 27 most important variables from all models. Variables are for the time period 2012 – 2019. Records with missing mortality status were excluded.

| Variable | Mortality Status |  | p-value<sup>2</sup> |
| --- | --- | --- | --- |
|  | 0, N = 220,829<sup>1</sup> | 1, N = 6,258<sup>1</sup> |  |
| **Age (years), mean (SD)** | 67.53 (11.23) | 70.77 (11.42) | <0.001 |
| **Female gender, n (%)** | 59,467 (27%) | 2,328 (37%) | <0.001 |
| **Body Mass Index, mean (SD)** | 28.49 (5.27) | 27.93 (5.49) | <0.001 |
| **Weight, mean (SD)** | 82.42 (16.75) | 78.98 (17.79) | <0.001 |
| **Number of previous operations, n (%)** |  |  | <0.001 |
| 0 | 212,318 (96%) | 5,288 (84%) |  |
| 1 | 7,600 (3.4%) | 829 (13%) |  |
| 2 | 775 (0.4%) | 105 (1.7%) |  |
| 3 | 115 (<0.1%) | 26 (0.4%) |  |
| 4 | 19 (<0.1%) | 9 (0.1%) |  |
| 5 | 1 (<0.1%) | 0 (0%) |  |
| 6 | 1 (<0.1%) | 1 (<0.1%) |  |
| **Urgency, n (%)** |  |  | <0.001 |
| 0 - Elective | 141,617 (64%) | 2,442 (39%) |  |
| 1 - Urgent | 72,090 (33%) | 2,134 (34%) |  |
| 2 - Emergency | 6,533 (3.0%) | 1,230 (20%) |  |
| 3 - Salvage | 589 (0.3%) | 452 (7.2%) |  |
| **First Operator Grade, n (%)** |  |  | <0.001 |
| 0 - Consultant | 179,959 (81%) | 5,729 (92%) |  |
| 1 - Associate specialist | 6,726 (3.0%) | 75 (1.2%) |  |
| 2 - Registrar/ SpR | 30,113 (14%) | 331 (5.3%) |  |
| 3 - SHO | 4,031 (1.8%) | 123 (2.0%) |  |

| Variable | Mortality Status |  |  |
| --- | --- | --- | --- |
|  | 0, N = 220,829<sup>1</sup> | 1, N = 6,258<sup>1</sup> | p-value<sup>2</sup> |
| **Critical Preoperative State, n (%)** | 7,255 (3.3%) | 1,382 (22%) | <0.001 |
| **Chronic pulmonary disease, n (%)** | 26,644 (12%) | 1,211 (19%) | <0.001 |
| **Extra-cardiac Arteriopathy, n (%)** | 22,327 (10%) | 1,215 (19%) | <0.001 |
| **Number of Grafts, n (%)** |  |  | <0.001 |
| 0 | 78,842 (36%) | 2,935 (47%) |  |
| 1 | 18,320 (8.3%) | 763 (12%) |  |
| 2 | 34,429 (16%) | 880 (14%) |  |
| 3 | 60,648 (27%) | 1,207 (19%) |  |
| 4 | 24,965 (11%) | 397 (6.3%) |  |
| 5 | 3,294 (1.5%) | 68 (1.1%) |  |
| 6 | 331 (0.1%) | 8 (0.1%) |  |
| **NYHA, n (%)** |  |  | <0.001 |
| 0 – I | 48,625 (22%) | 1,055 (17%) |  |
| 1 – II | 96,888 (44%) | 1,609 (26%) |  |
| 2 – III | 64,049 (29%) | 2,228 (36%) |  |
| 3 – IV | 11,267 (5.1%) | 1,366 (22%) |  |
| **Creatinine, mean (SD)** | 92.61 (47.15) | 119.15 (84.48) | <0.001 |
| **Cardiac Rhythm, n (%)** |  |  | <0.001 |
| 0 – Sinus | 193,158 (87%) | 4,677 (75%) |  |
| 1 – Preop AF | 24,240 (11%) | 1,309 (21%) |  |
| 2 – Preop VFT | 399 (0.2%) | 60 (1.0%) |  |
| 3 – Preop CHB or pacing | 3,032 (1.4%) | 212 (3.4%) |  |
| **Number of Valves, n (%)** |  |  | <0.001 |
| 0 | 133,133 (60%) | 2,941 (47%) |  |
| 1 | 77,621 (35%) | 2,545 (41%) |  |
| 2 | 9,148 (4.1%) | 659 (11%) |  |
| 3 | 916 (0.4%) | 111 (1.8%) |  |
| 4 | 11 (<0.1%) | 2 (<0.1%) |  |
| **Pulmonary Artery Systolic Pressure, mean (SD)** | 23.65 (11.87) | 27.88 (16.80) | <0.001 |
| **MV Procedure, n (%)** |  |  | <0.001 |
| 0 - None | 191,803 (87%) | 4,870 (78%) |  |
| 1 - Repair | 17,633 (8.0%) | 467 (7.5%) |  |

| Mortality Status |  |  |  |
| --- | --- | --- | --- |
| Variable | 0, N = 220,829<sup>1</sup> | 1, N = 6,258<sup>1</sup> | p-value<sup>2</sup> |
| 2 - Replacement | 11,393 (5.2%) | 921 (15%) |  |
| **Number of previous Myocardial infarction, n (%)** |  |  | <0.001 |
| 0 | 150,612 (68%) | 3,916 (63%) |  |
| 1 | 60,266 (27%) | 1,887 (30%) |  |
| 2 | 9,951 (4.5%) | 455 (7.3%) |  |
| **Ascending aorta procedure, n (%)** | 8,011 (3.6%) | 769 (12%) | <0.001 |
| **Previous valve surgery, n (%)** | 5,701 (2.6%) | 669 (11%) | <0.001 |
| **Cardiogenic Shock, n (%)** | 2,379 (1.1%) | 840 (13%) | <0.001 |
| **Pre-op Dialysis, n (%)** |  |  | <0.001 |
| 0 - None | 217,789 (99%) | 5,764 (92%) |  |
| 1 - No dialysis but AKI | 853 (0.4%) | 164 (2.6%) |  |
| 2 - AKI within 6wks of surgery needing dialysis | 696 (0.3%) | 157 (2.5%) |  |
| 3 - CKD dialysis | 1,491 (0.7%) | 173 (2.8%) |  |
| **Days between LHC and cardiac surgery date, mean (SD)** | 91.07 (858.72) | 95.97 (468.56) | <0.001 |
| **LVEF, n (%)** | 54.01 (7.38) | 51.43 (9.78) | <0.001 |
| **Inotropes, n (%)** | 2,114 (1.0%) | 726 (12%) | <0.001 |
| **Aortic arch procedure, n (%)** | 1,286 (0.6%) | 216 (3.5%) | <0.001 |
| **Aortic root procedure, n (%)** | 4,069 (1.8%) | 387 (6.2%) | <0.001 |

<sup>1</sup>Mean (SD) or Frequency (%)
<sup>2</sup>Wilcoxon rank sum test; Pearson's Chi-squared test; Fisher's exact test
AF - Atrial Fibrillation; VFT - Ventricular fibrillation or ventricular tachycardia; CHB - Complete heart block / paced; CKD - Chronic kidney disease; LHC - Left Heart Catheterization; LVEF - Left ventricular ejection fraction.

### Baseline variable characteristics

Following data pre-processing, there were no missing values (Figure S4). Correlation analysis of all 61 variables considered for the non-variable selected models showed that there was no concern for multi-collinearity (Figure S3). After pre-processing and pooling 18 most important variables from each model and retaining only unique variables, 27 variables were found to have strong evidence of being associated with outcome (Table 1, p <0.001).

### Variable Importance Characteristics

The top 18 important variables for each of the non-variable selected models are shown in Table S12. Detailed importance scores for the variables of each model are shown for Mixed Effects Xgboost: Table S4, Figure S5, S6; MERF: Table S5, Figure S7, S8; Xgboost – Centre: Table S6, Figure S9, S10; Random Forest (RF) – Centre: Table S7, Figure S11, S12; Random Forest + Centre (hot-encoded): Table S8, Figure S13, S14; Xgboost + Centre (hot-encoded): Table S9, Figure S15, S16; GPBoost + Centre: Table S10, Figure S17 and S18. It can be seen that models of the same general type without a centre and that with a centre (hot-encoded) have a more similar ranking of important variables than the corresponding mixed-effects model. Urgency has the highest frequency of being selected as the most important predictor of mortality across models, followed by Age, Creatinine, NYHA, Previous Surgery, Pulmonary Artery Systolic Pressure and Weight.

### Hyperparameter Tuning

The optimal hyperparameters for non-variable selected and variable selected models are shown in Table S2 and S3, respectively.

## Assessment of model performance

### Non-variable selected model

#### Adapted Rain plots

For non-variable selected models, it can be seen in Figure 2a that AUC performance is generally consistent across models with GPBoost having slightly lower AUC than other models. Mixed-effects RF + RE: centre and Mixed Effects Xgboost + RE: centre models performed poorly compared to other models across several metrics, namely: adjusted ECE, adjusted Brier, and net benefit. Apart from these two models, the other models performed comparably in terms of net benefit. These two models achieved higher F1 scores than GPBoost but were outperformed by all remaining models. Xgboost + Centre (hot-encoded) demonstrated the highest overall performance in terms of magnitude and ranking as shown in a detailed report of individual metric results comprising the CEM (Table 2).

**Figure 2. a).**
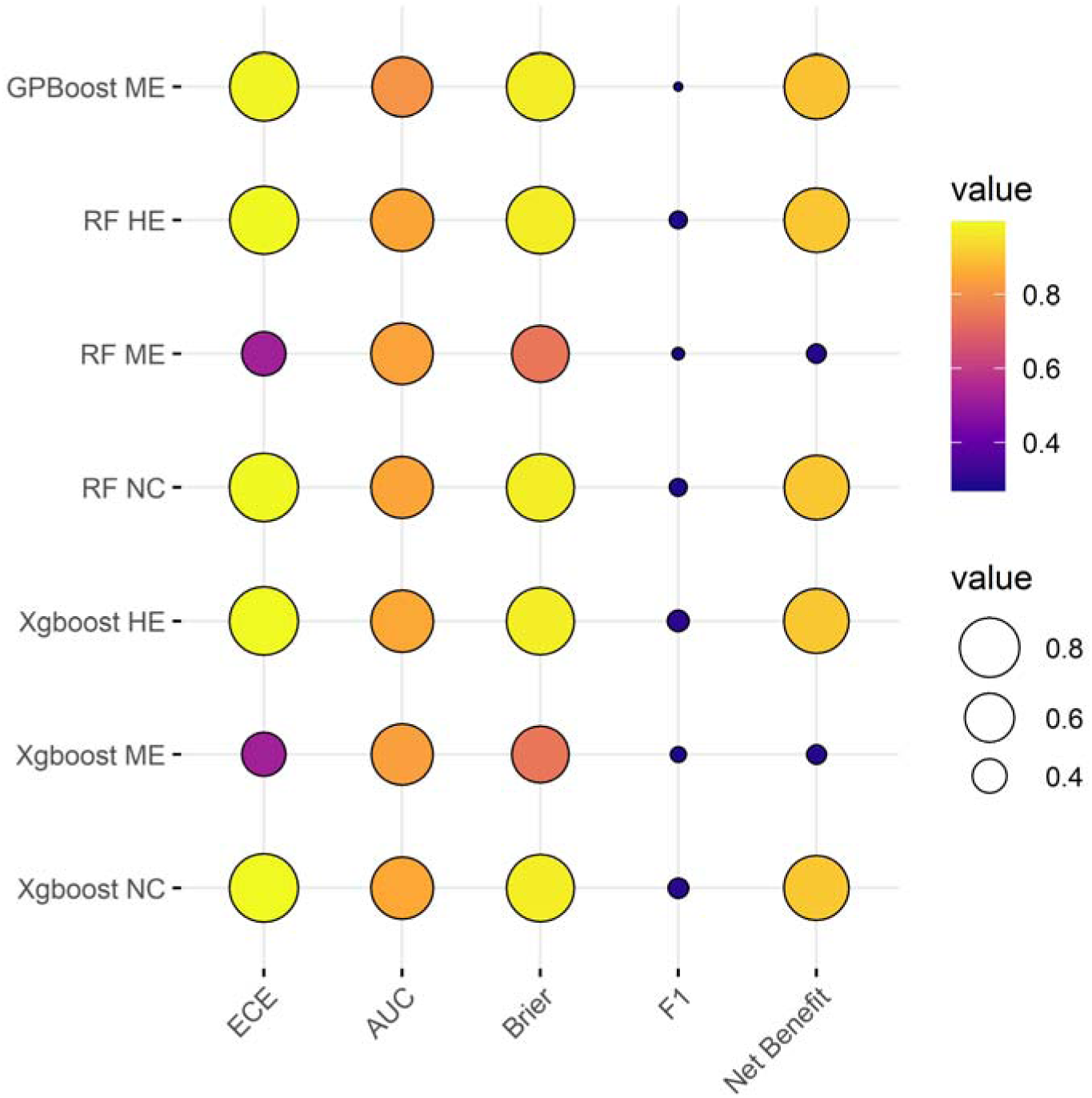
Non variable selected models: adapted Rain plot of CEM constituent metrics by model; larger sized spheres represent higher metric performance and vice versa.

**Table 2.** Non-variable selected models: Geometric Mean of Individual metrics; CEM refs to Clinical Effective Metric; Standard deviation and 95% CI are shown for CEM; adjusted 1 – ECE and 1 – Brier score values are shown; net benefit is average absolute overall benefit across all thresholds.

| Model Category | ECE | AUC | Brier | F1 | Net Benefit | CEM Mean | CEM S.D | CEM CI Lower limit | CEM CI Upper limit |
| --- | --- | --- | --- | --- | --- | --- | --- | --- | --- |
| GPBoost ME | 0.991 | 0.808 | 0.976 | 0.267 | 0.899 | 0.716 | 0.005 | 0.715 | 0.716 |
| RF HE | 0.995 | 0.849 | 0.977 | 0.284 | 0.906 | 0.734 | 0.005 | 0.733 | 0.734 |
| RF ME | 0.520 | 0.843 | 0.744 | 0.270 | 0.290 | 0.480 | 0.004 | 0.480 | 0.481 |
| RF NC | 0.996 | 0.848 | 0.977 | 0.284 | 0.906 | 0.734 | 0.005 | 0.733 | 0.734 |
| Xgboost HE | 0.997 | 0.855 | 0.977 | 0.300 | 0.908 | 0.743 | 0.005 | 0.743 | 0.744 |
| Xgboost ME | 0.520 | 0.835 | 0.744 | 0.278 | 0.291 | 0.483 | 0.003 | 0.482 | 0.483 |
| Xgboost NC | 0.997 | 0.854 | 0.977 | 0.296 | 0.908 | 0.741 | 0.005 | 0.741 | 0.741 |

#### Statistical Analyses

No extreme outliers were found. The CEM scores was normally distributed for all models, as assessed by Shapiro-Wilk’s test (p > 0.05). There was strong evidence of a difference across models p < 0.0001, except between RF + Centre (hot-encoded) and RF – Centre (Table S11 and Figure S19). Dunnett’s test showed that there was strong evidence that Xgboost + Centre (hot-encoded) was superior to the Xgboost – Centre model (p < 2e-16, Table 3). There was strong evidence that Xgboost – Centre model outperformed all other models, with Mixed effects RF + RE: centre performing worst, followed by Mixed Effects Xgboost + RE: centre (CEM difference: –0.2605 and –0.2583, respectively). RF – Centre and RF + Centre (hot-encoded) had similar performance (CEM difference: –0.0073) and have performance rankings immediately below Xgboost – Centre, but above GPBoost.

**Table 3.** Non-variable selected models: Dunnett’s test with Xgboost – Centre as control; 95% family-wise confidence level are shown as well as mean difference in CEM and p-values; NC: no centre; HE: hot-encoded centre; ME: mixed effects.

| Group 1 | Group 2 | CEM Difference (1-2) | P Value | 95% CI |  |
| --- | --- | --- | --- | --- | --- |
|  |  |  |  | Lower Bound | Upper Bound |
| GPBoost ME | Xgboost NC (Control) | -0.0252 | <2e-16 *** | -0.0257 | -0.0247 |
| RF HE |  | -0.0073 | <2e-16 *** | -0.0078 | -0.0067 |
| RF ME |  | -0.2605 | <2e-16 *** | -0.2610 | -0.2600 |
| RF NC |  | -0.0073 | <2e-16 *** | -0.0078 | -0.0068 |
| Xgboost HE |  | 0.0024 | <2e-16 *** | 0.0019 | 0.0029 |
| Xgboost ME |  | -0.2583 | <2e-16 *** | -0.2589 | -0.2578 |

### Variable selected Xgboost

#### Adapted Rain plots

Model performance differences showed a similar overall pattern for variable selected compared to non-variable selected models (Figure S21). GPBoost showed lower AUC compared to other models, with the contrast in difference being greater for variable selected than for non-variable selected models. There was lower variation in F1 score for variable selected models, with all models outperforming GPBoost. Although mixed-effects models RF and Xgboost obtained a higher net benefit for variable selected models, the net benefit performance was markedly lower compared to other models than for non-variable selected models. A detailed report of individual metric results comprising the CEM is given in (Table S15).

#### Statistical Analyses

No extreme outliers were found. The CEM scores were normally distributed for all models, as assessed by Shapiro-Wilk’s test (p > 0.05). There was strong evidence of a difference across models p < 0.0001, except between: a) RF + Centre (hot-encoded) and RF – Centre; b) Mixed-effects RF and Mixed Effects Xgboost; c) Xgboost + Centre (hot-encoded) and Xgboost – Centre (Table S13 and Figure S20). Dunnett’s test showed that there is minute but insignificant increase in performance between Xgboost + Centre (hot-encoded) and Xgboost – Centre model (CEM Difference: 0.0002, p = 0.94, Table S14). Xgboost – Centre significantly outperformed all other models except Xgboost + Centre (hot-encoded). The next best performing model was RF + Centre (hot-encoded) followed by RF – Centre, GPBoost and Mixed-effects RF. Mixed-effects Xgboost demonstrated the worst performance (CEM Difference: –0.2539).

### Highest performing: non-variable vs. variable selected model vs. Xgboost (ES II variables)

Xgboost + Centre (hot-encoded; 102 variables), Xgboost + Centre (hot-encoded; 18 variables) and Xgboost (ES II) variables models were compared. No extreme outliers were found. The CEM scores was normally distributed for all three models except Xgboost (ES II), as assessed by Shapiro-Wilk’s test (p > 0.05). A histogram plot of the Xgboost (ES II) CEM values did not show substantial deviation from the normal distribution. There was strong evidence of a difference across models p < 0.0001 (Table S16 and Figure 2b).

**Figure 2. b).**
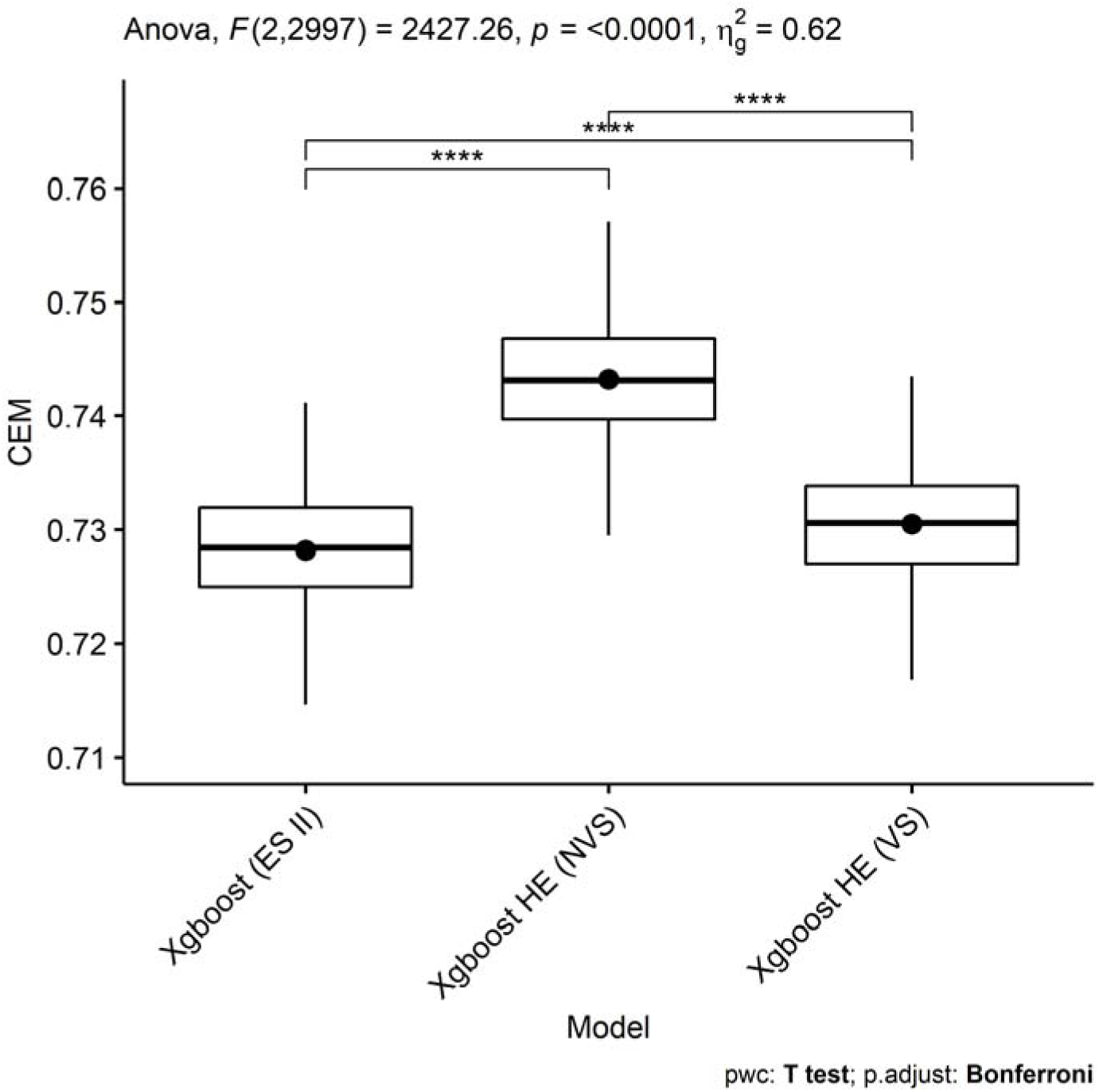
CEM performances of Xgboost (ES II), the best non-variable and variable selected models are compared using multiple pairwise paired t-tests with Bonferroni correction; NVS: non-variable selected; VS: variable selected; ES II: EuroSCORE II.

Dunnett’s test showed that there was strong evidence that both non-variables selected (NVS) and variables selected (VS) Xgboost + Centre (hot-encoded) models were superior to the Xgboost (ES II) model (p < 6.3e-15, Table S17), with NVS Xgboost + Centre (hot-encoded) model having the best performance, followed by the VS Xgboost + Centre (hot-encoded) model (CEM Difference: 0.0150 and 0.0023, respectively).

### Bayesian Networks

The Bayesian network for the interactions between the top 18 important variables, from the optimal model: Xgboost + Centre (hot-encoded), and the outcome (mtly) shows that Urgency, Age, Critical Preoperative State (CPS), NHYA, Number of Valves, Number of previous operations (PrevOp) have direct relationship / path to the outcome variable (Figure 3). CPS confounds the relationship between NYHA and the outcome; Age confounds the relationship between Urgency and the outcome; NYHA confounds the relationship between Number of Valves and the outcome; Urgency confounds the relationship between CPS and the outcome; Number of Valves and Age both confounds the relationship between PrevOp and the outcome. Creatinine is a potential collider confounder that could bias the relationships between Urgency and the outcome.[39]

**Figure 3.**
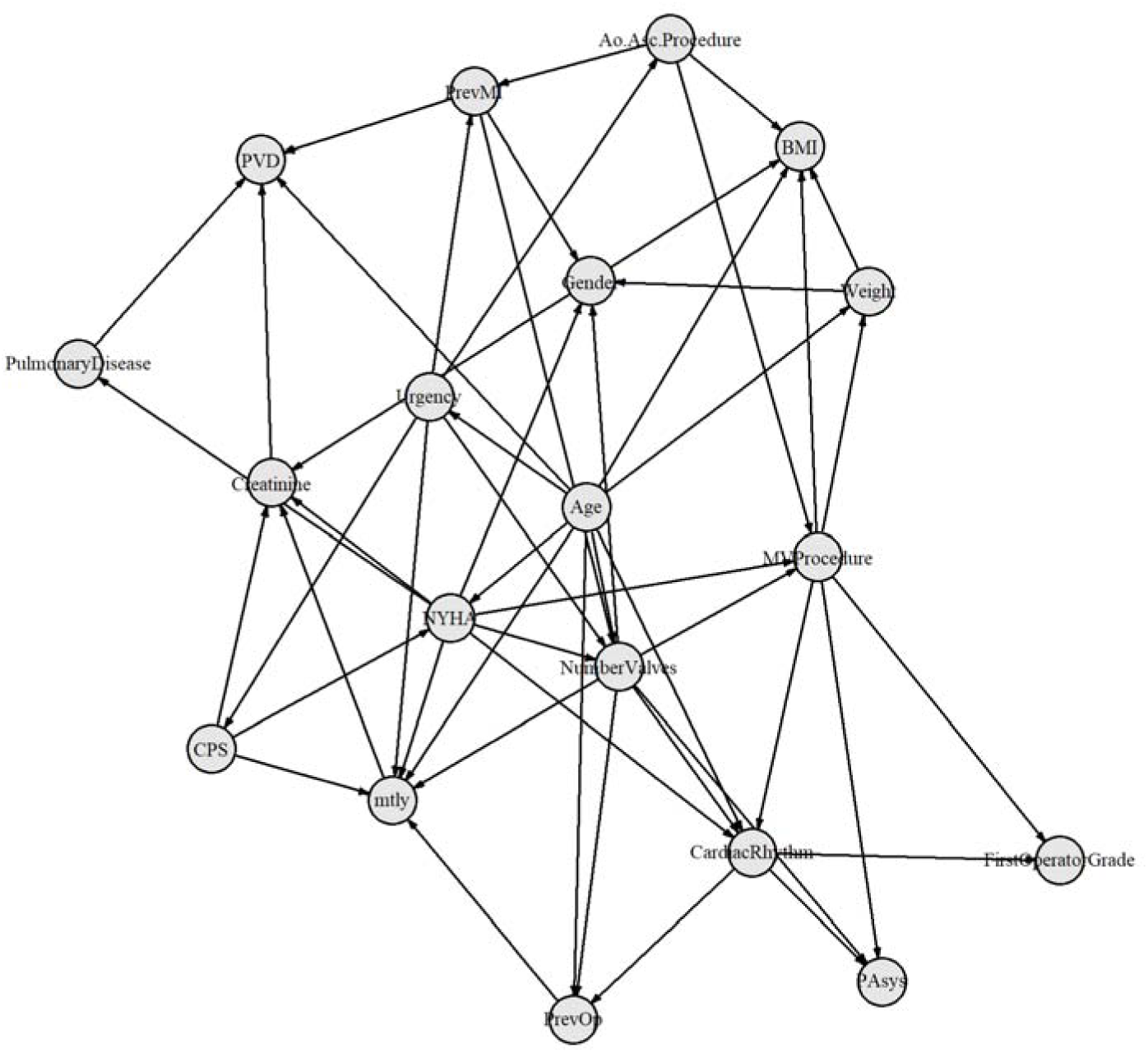
Bayesian network of interactions between the top 18 important variables, from Xgboost + Centre (hot-encoded) model, and the outcome, i.e. mtly: 0 – survival, 1 – non survival.

The Bayesian network for the interactions between all the variables, from the optimal model: Xgboost + Centre (hot-encoded), and the outcome (mtly) shows that only Urgency and Cardiogenic Shock have a direct relationship/path to the outcome (Figure 4). Urgency confounds the relationship between Cardiogenic Shock and the outcome. Urgency mediates the relationship between Active Endocarditis (Endocarditis) and the outcome, whilst Endocarditis mediates the relationship between Mitral Valve procedure (MVProcedure) and Urgency. Urgency mediates the relationship between “Interval between surgery and myocardial infarction (MI)” (IntervalMI) and the outcome. Urgency mediates the relationship between ascending aorta procedure (Ao.Asc.Procedure) and the outcome. Ao.Asc.Procedure mediates the relationship between Aortic root procedure (Ao.Root.Procedure) and Urgency.

**Figure 4.**
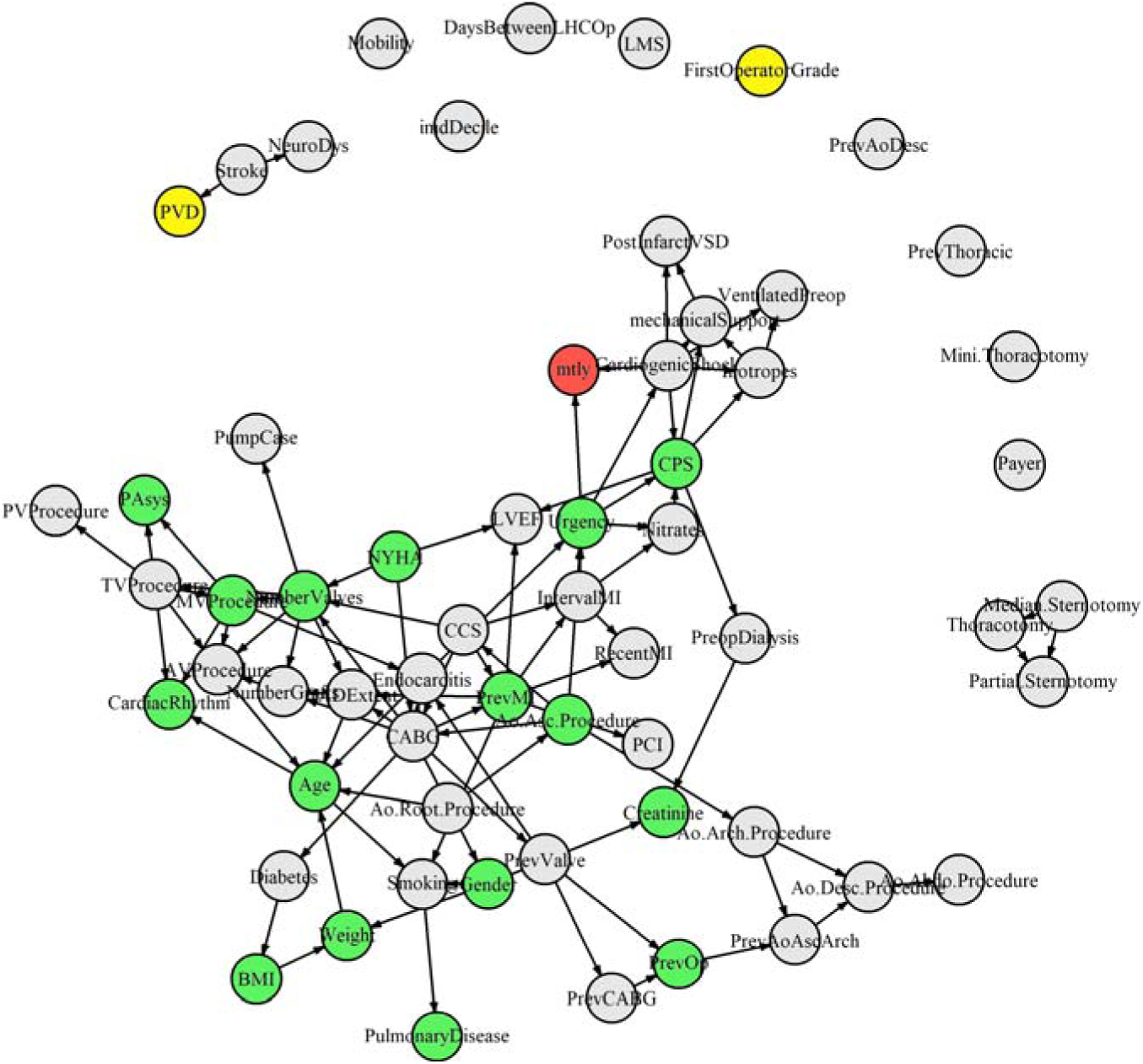
Bayesian network of interactions between all variables, from the non-variable selected Xgboost + Centre (hot-encoded) model, and the outcome (red), i.e. mtly: 0 – survival, 1 – non survival. The top 18 important variables are shown in green: within main network or in yellow: outside of the main network.

## DISCUSSION

ML approaches have the advantage of not be limited by linear statistical assumptions, and the number of clinical variables can theoretically increase exponentially. In addition, the variations and residual confounding that historically hindered the usefulness of cardiac surgery risk stratification scores can potentially be taken into account. However, these potential goldmines in clinical data usage have yet to be fully harvested. One example of this problem is exemplified in the controversial decision over whether to use a single procedural or a multi-procedural (universal) risk score/model.

While some studies have investigated the potential to devise scoring systems for specific surgical procedures such as tricuspid valve surgery using additional scores for other diseases such as a score for end-stage liver disease (MELD),[40] they have been limited by samples size of cohorts, unavailability of multi-centre data and limited use of holdout data. Conversely, universal (or general) risk scores have been developed focusing on a wider range of procedures than that by ES II and STS-PROM,[41] such as the American College of Surgeons National Surgical Quality Improvement Program (ACS-NSQIP) for mortality and morbidity for more than 100 different procedures.[42] Universal models such as the EuroSCORE and ES II have the benefit of allowing a large number of covariates to be included to improve model performance for low event rate (e.g. <1%) cardiac surgery datasets,[41] as well as allowing surgeons to evaluate the risk for nearly any combination of cardiac procedures, something that the procedure specific STS-PROM score does not allow.[41] There is mixed evidence as to whether a universal or procedure-specific score is preferable.[43]

Unlike our study, the current state of procedural specific and universal scores described above did not adjusted for hospital-based variation using a machine learning-based approach. This study also provides evidence for the use of larger number of covariates in universal cardiac surgery risk prediction models, and substantially increases the number of risk factors compared to only 18 variables of ES II. This was achieved by comparing ML models built from a combination of a large set of 61 variables, and adjusting for geographical variation as a result of 42 different hospital contributions. In addition, by using a larger multi-centre national dataset, the number of events (6,258; mortality rate of 2.76%) increased compared to smaller sized studies (<1%).[41] Therefore, despite using a substantially larger number of variables in the best performing Xgboost non-variable selected model with 102 variables, the Events Per Variable (EPV) rate remained (EPV = 6,258 / 102 = 61) substantially larger than that previously found through simulation to be necessary for low bias and error in prediction modelling (EPV > 25).[13]

This study provides strong evidence that the ES II variables are suboptimal for risk stratification modelling and that a larger set of 60 variables with additional adjustment for hospital variation provides superior performance to ML models built using ES II variables. The Xgboost model with hospital location adjustment of the large variable set performed best overall. Whilst the parsimonious models using the small (18) number of variables provided significiantly weaker performance than the full variable set model, we demonstrated that there was significant evidence that the new small (18) variable models (Xgboost variations) outperformed the ES II 18 variable model (Xgboost). However, the improvements through adjustment for hospital location in the small variable set is so small through the dummy coding approach that this may not be necessary.

Notably, this study provides strong evidence that adjustment for hospital location improves ML model performance, but only in the presence of adjustment using a large set of variables and that cardiac surgery risk models with relatively small number of variables may not require this adjustment process. One possible reason for this is that in the small variable set models, more confounders are not being adjusted for. It is also worth noting that this effect applied to certain ML models, namely Xgboost, but did not apply to Random Forest. This work also shows that adjustment, whereby hospital location variables are separated into individual binary or dummy variables, demonstrated strong evidence of being superior over adjusting for location using a single vector random variable in a mixed-effects model. It is possible that separating out the hospital random effects into a multi-dimension set of individual vectors enabled the non-linear machine learning models to better adjust for the complex interactions across differences (or heterogeneity) in patient and operative characteristics across different cardiac centres.

One limitation of universal scores is that unadjusted variables that are not part of the score may negatively impact procedures that are more reliant on such variables, than other procedures, leading to inconsistency in procedural performances.[44] It has been highlighted that the high importance of certain interventional variables in risk scores is confounded by other factors, especially for the sickest patients,[11] making scores such as ES II and STS less useful for procedures for which confounding variables are not included.

This study is one of the first in its kind to demonstrate the importance of using Bayesian Networks (BN) for confounding analysis in the clinical setting as an alternative to the typically used Propensity score, which although useful, may overlook subtle causal relationships that could confound the real relationship between clinical variables and the outcome. The proposed BN approach enables clinicians to interpret the causal relationships in light of the confounding and is well suited to studies involving large variables such as the current study. One interesting observation is that by including the full set of variables in contrast to only the top 18 most important variables, many of the previously identified confounding relationships across variables disappeared. We identified that Cardiogenic Shock is the main cause of mortality following cardiac surgery, even though causation does not necessarily indicate optimal importance for prediction.

Although Urgency (or highly urgent) procedures are also more likely to result in a higher risk of mortality, it could be seen that MV procedures for Endocarditis are likely to result in higher Urgency cases and consequently lead to a higher risk of surgical mortality. Another interesting finding is that the longer the interval between myocardial infarction (MI) and surgery, the higher the Urgency and consequently the higher the mortality risk. Clinical efforts should therefore target minimsing the interval between MI and surgery. We also identified ascending aorta and specifically root procedure as high risk.

This study also provides a novel application of the adapted Rain plot for taking a wholistic view of the individual constituent metrics within the Clinical Effective Metric (CEM) consensus metric, and enables the filtering of high performance models. This provides a useful tool for clinicans to better understand how the CEM arrives at its ranking of competitive risk scores.

## Limitations and Future work

This study is not without limitations. As the STS-PROM model coefficients are not made publicly available, we were not able to compare our universal model against a procedural specific model using this combination of new variables. Future studies should aim to compare the effects of pre-selecting features[18] using linear mixed effects adjustement of hospital variations in relation to the hot-encoding adjustment of random effects as well as considering multiple random effects including adjustment for surgeon differences.[17] To validate the findings of this study, future studies may also compare the models considered herewithin using the linear mixed effects adjustment as a post-processing step.[15,16] Although the optimal models proposed in this study outperformed all models from our previous studies,[21,22] one possible reason for the mixed-effects models underperforming compared to other models is that mixed-effects ML models are typically better suited to regression tasks and are less well suited to classification tasks. Although the current study is mainly focused on the generalisability in terms of performance ranking on the hold out dataset, future work should investigate the effects of overfitting by considering approaches such as using, for example, the normalised ratio of holdout to training performance metric values. While a large number of clinically relevant variables have been taken into account, there may still be variables and hence residual confounding not adjusted for due to the availability of variables in the dataset. The effects of including or excluding variables such as Creatinine that demonstrated potential collider confounding relationships need to be further assessed.

Further work is required to enhance the performance of mixed-effects models for the purpose of cardiac surgery risk classification.

## CONCLUSION

This study based on a multi-centre national registry dataset comprising 42 UK hospitals highlights the importance of a larger set of potential confounding variables previously not considered by the EuroSCORE. II. Furthermore, it suggests to adjust for hospital variation with specific recommendations for applying the ML model to study big data whereby each hospital is separated into individual binary input variables. We identified an ML-based hospital variation adjusted risk score comprising a large number of clinically predictive variables that increases risk stratification performance on hold out dataset, removing the need to perform variable selection and reduction. We demonstrated the concept of Bayesian Networks for cardiac surgery mortality associated causal relationship analysis following comparative risk score selection and identified Cardiogenic Shock, Urgency, the interval between myocardial infarction (MI) and surgery, MV procedures for Endocarditis, ascending aorta procedures and aortic root procedures to either directly or indirectly cause a higher risk of cardiac surgery mortality. Lastly, this study highlights the versatility of the adapted Rain plot for rapid clinical assessment of which risk score should be considered for cardiac surgery patient selection across multiple consistuent metrics and risk scores. It is recommended that this approach should be used in conjunction with the more robust CEM consensus metric and the Bayesian Network whereby possible. Future work will examine this new scoring approach in the context of performance drift and take into account procedural adjustments.

## Funding

This work was supported by a grant from the BHF-Turing Institute and the NIHR Biomedical Research Centre at University Hospitals Bristol and Weston NHS Foundation Trust and the University of Bristol.

## Contributorship

T.D., S.S., D.P.F., J.C., B.Z., P.N., M.C, U.B., A.J., A.D., G.D.A. contributed to experimental design. T.D. and S.S. acquired data. T.D. and S.S performed the data preprocessing. T.D. wrote the source code to perform the experiments, and are accountable for all aspects of the work. T.D., S.S, D.P.F., J.C., B.Z., P.N., M.C., A.J., A.D., G.D.A analyzed the results. T.D. wrote the first version of the paper. All authors revised the paper and approve the submission.

## Data availability

All data used in this study are from the National Adult Cardiac Surgery Audit (NACSA) dataset. These data may be requested from Healthcare Quality Improvement Partnership (HQIP), https://www.hqip.org.uk/national-programmes/accessing-ncapop-data/#.Ys6gN-zMLdp. Code for deriving training, update, and hold-out datasets is available on GitHub and authors can provide confirmatory de-identified record IDs for each set upon reasonable request.

## Competing Interests

All authors declare that there are no competing interests.

## Ethics statement

The study was approved by the Health Research Authority (HRA) and Health and Care Research Wales (HCRW) in 23 of July 2019, IRAS project ID: 278171 and a waiver for patients’ consent was obtained.

Guarantor TD.

## Code availability

All source code used in this study are available on GitHub (https://github.com/s0810110/EnsembleScoreAdaption). Analyses were performed using Scikit-learn v0.23.1, Keras v2.4.0, STATA-MP version 17 and R v4.0.2.

## Notes

### Competing Interest Statement

The authors have declared no competing interest.

### Author Declarations

The register-based cohort study is part of a research approved by the Health Research Authority (HRA) and Health and Care Research Wales and a waiver for patients' consent was waived (HCRW) (IRAS ID: 278171).

