## Supplementary materials for "Random effects adjustment in machine learning models for cardiac surgery risk prediction: a benchmarking study"

*Tim Dong MSc, Shubhra Sinha MD, Daniel P Fudulu, PhD, Jeremy Chan MD, Ben Zhai PhD, Pradeep Narayan FRCS(CTh), Massimo Caputo MD, Andy Judge PhD, Arnaldo Dimagli MD, Umberto Benedetto PhD and Gianni D. Angelini MD.*

**Abbreviations and Acronyms:**

AUC area under receiver operating characteristic curve;

CEM Clinical Effective Metric;

ECE Expected Calibration Error;

ES II Euroscore II;

AI Artificial intelligence;

ML machine learning;

RF random forest;

XGBoost extreme gradient boosted trees

SHAP (SHapley Additive exPlanations)

NVS non-variable selected

VS variable selected

Table S1. 61 Initial set of variables

| **Count** | **Id** | **Name** | **Number of Levels** | **Present in Other Scores** | **Data Transformation** | **Handling of missing data** |
| --- | --- | --- | --- | --- | --- | --- |
| 1 | 5 | HospCode | Categorical |  | Two dataset versions:  1.convert to ordinal  2.hot-encode | No missing data |
| 2 | 7 | Gender | 0.Male 1.Female | LogES/ESII/ SYNTAX II | Same | Set NAs to 0 |
| 3 | 8 | Payer | 1.NHS, 2.Private |  | 0.Private, 1.NHS  Same.[1] | Set NAs as 1 (1 after transformation) |
| 4 | 9 | CCS | 0-4 | ESII | Same | Set NAs to 0 |
| 5 | 10 | NYHA | 1-4 | ESII | 0-3 | Set NAs to 1 |
| 6 | 11 | PrevMI | 0.None 1.One 2.Two | LogES/ESII | Same | Set NAs to 0 |
| 7 | 12 | IntervalMI | 0. No 1. 6hrs 2. 6-24hrs 3. 1-30days 4. 31-90 |  | 0. No  1. 31-90 days  2. 1-30days  3. 6-24hrs  4. 6hrs | If PrevMI=0 then change to 0.  Set NAs to 0. |
| 8 | 13 | PCI | 0.No  1.<24hrs,same admission  2.>24hrs,same admission  3.>24hrs,prev admission |  | 0.No  1.>24hrs,prev admission  2.>24hrs,same admission  3.<24hrs,same admission | Set NAs to 0 |
| 9 | 15 | PrevCABG | 0.No 1. Yes |  | Same | Set NAs to 0 |
| 10 | 16 | PrevValve | 0.No 1. Yes |  | Same | Set NAs to 0 |
| 11 | 19 | PrevAoAscArch | 0.No 1. Yes |  | Same | Set NAs to 0 |
| 12 | 20 | PrevAoDesc | 0.No 1. Yes |  | Same | Set NAs to 0 |
| 13 | 21 | PrevThoracic | 0.No 1. Yes |  | Same | Set NAs to 0 |
| 14 | 30 | Diabetes | 0.No 1.Diet 2.Oral 3.Insulin | LogES/ESII | Same | Set NAs to 0 |
| 15 | 32 | Smoking | 0.Never 1.Ex 2.Current |  | Same | Set NAs to 0 |
| 16 | 33 | PulmonaryDisease | 0.None 1.COPD with inhalers  2.Asthma | LogES/ESII/ SYNTAX II | set both 1 and 2 to 1, PulmonaryDisease:  0.No 1.Yes[2] | Set NAs to 0 |
| 17 | 34 | Stroke | 0.None 1.TIA 2.CVA with recovery 3.CVA with deficit |  | same | Set NAs to 0 |
| 18 | 35 | NeuroDys | 0.No 1.Yes | Use mobility for ESII | Same | Set NAs to 0 |
| 19 | 36 | PVD | 0.No 1.Yes | LogES/ESII/ SYNTAX II | Same | If PrevPVD=1 then change to 1  Set NAs as 0 |
| 20 | 38 | - CardiacRhythm   - 0.Sinus   - 1.PreopAF   - 2.PreopVFT   - 3.PreopCHB.pacing | 0.No 1.Yes |  | Combine 1,2,3 into one variable with levels 1,2,3; Set all remaining zeros in new variable as 0.Sinus Rhythm:  0.Sinus  1.PreopAF  2.PreopVFT  3.PreopCHB.pacing | Set NAs as 0 |
| 21 | 46 | LMS (left main stem disease) | 0.None 1.LMS>50% | SYNTAX II | Same | If LMS=1 then CADExtent=2 or 3;  Set NAs as 0 |
| 22 | 47 | PAsys | Numeric |  | Same | Impute using median |
| 23 | 50 | LVEF | Numeric | SYNTAX II | Same | Impute using median |
| 24 | 55 | CardiogenicShock | 0.No 1. Yes |  | Same | Set NAs as 0 |
| 25 | 57 | VentilatedPreop | 0.No 1. Yes |  | Same | Set NAs as 0 |
| 26 | 58 | Urgency | 1.Elective 2.Urgent 3.Emergency 4. Salvage | LogES/ESII | 0.Elective 1.Urgent 2.Emergency 3. Salvage | Set NAs as 1 (0 after transformation) |
| 27 | 59 | PrevOp | Numerical – no. of prev operations |  | Same | If >1 then set PrevSurg(col 14)=1  Set NAs as 0 |
| 28 | 62 | BMI | Derived from height and weight |  | Same[3] | Impute using median |
| 29 | 63 | Mobility(Poor Mobility) | 0.No 1.Yes |  | Same | Set NAs to 0 |
| 30 | 69 | FirstOperatorGrade | 1. Consultant 2. Associate specialist 3. Registrar/ SpR 4. SHO | - | 0.Consultant  1.Associate specialist  2.Registrar/ SpR  3.SHO | Set NAs to 0 |
| 31 | 95 | CABG | 0.No 1.Yes |  | Same | If col97 or col240>1 then 1.  Set NAs to 0 |
| 32 | 98 | NumberValves | Numerical |  | Same | If >1 then change col96 to 1  Set NAs to 0 |
| 33 | 106 | AVProcedure | 0.None  1.Replacement  2.Repair  3.Repair with ring  4.Repair w/o ring  5.Commisurotomy  6.Excision  7.Inspection |  | Set 7 as 0; re-group as:  0.None  1.Repair (combine 2-6)  2.Replacement | If 1 then change col96 to 1  Set NAs to 0 |
| 34 | 113 | PreopDialysis | 0.None  1.AKI within 6wks of surgery needing dialysis  2.CKD^a^ dialysis  3.No dialysis but AKI  ^a^Chronic kidney disease |  | 0.None  1.No dialysis but AKI  2.AKI within 6wks of surgery needing dialysis  3.CKD dialysis | Set NAs to 0 |
| 35 | 114 | Creatinine | Numeric |  | Same | Outliers as per nacsa;  Impute using median |
| 36 | 118 | Age | Numerical | LogES/ESII | Same | Round up  No missing values identified |
| 37 | 136 | PumpCase | 0. Off pump  1. On pump |  | Same[4] | If CPB>0 then change to 1;  Set NAs to 0 |
| 38 | 181 | CPS | Derived  0.No 1.Yes | LogES/ESII | Same |  |
| 39 | 183 | Endocarditis | Derived  0.No 1.Yes | LogES/ESII | Same | Set NAs to 0 |
| 40 | 184 | RecentMI | Derived  0.No 1.Yes | LogES/ESII | Same | Set NAs to 0 |
| 41 | 185 | PostInfarctVSD | Derived  0.No 1.Yes | LogES | Same | Set NAs to 0 |
| 42 | 233 | MVProcedure | 0 none, 1 replacement, 2 repair, 3 repair with ring, 4 repair without ring, 5 isolated commisurotomy, 6 excision only, 7 inspection |  | Set 7 as 0; re-group as:  0.None  1.Repair (combine 2-6)  2.Replacement | Set NAs to 0 |
| 43 | 237 | TVProcedure | 0 none, 1 replacement, 2 repair, 3 repair with ring, 4 repair without ring, 5 isolated commisurotomy, 6 excision only, 7 inspection |  | Set 7 as 0; re-group as:  0.None  1.Repair (combine 2-6)  2.Replacement | Set NAs to 0 |
| 44 | 244 | PVProcedure | DaysBetweenLHCOp |  | Set 7 as 0; re-group as:  0.None  1.Repair (combine 2-6)  2.Replacement | Set NAs to 0 |
| 45 | 44 | DaysBetweenLHCOp | Derived from LHCDate and OpDate  Numerical |  | Same | Impute using median |
| 46 | 45 | CADExtent | 0.None>50 1.One>50 2.Two>50 3.Three>50  4.Not investigated |  | Weight | Set NA to 0 |
| 47 | 54 | Nitrates | 0.No 1.Yes |  | Same | Set NAs as 0 |
| 48 | 56 | Inotropes | 0.No 1.Yes |  | Same | Set NAs as 0 |
| 49 | 61 | Weight | Numerical |  | Same | Outliers as per NACSA |
| 50 | 97 | NumberGrafts | Numerical |  | Same | If >1 then change col95 to 1;  Remove if >6 (nacsa use 11 as a cut off);  Set NAs as 0 |
| 51 | 160 | Ao.Root.Procedure | 1.IPG+/-extension into the arch  2.IPG+reimplantation  3.Tube graft + sep. AVR  4.Bentall  5.VSRR  6.Homograft  7.Autograft/Ross  8. Aortic patch graft  9.SoV repair  10.Red aortoplasty |  | Set as 0 if none; 1 if any of 1-10 is true:  0.No 1.Yes | If 1 then col74 =1;  Set NAs as 0 |
| 52 | 161 | Ao.Asc.Procedure | 1.IPG+/-extension into the arch  2.IPG+reimplantation  3.Tube graft + sep. AVR  4.Bentall  5.VSRR  6.Homograft  7.Autograft/Ross  8. Aortic patch graft  9.SoV repair  10.Red aortoplasty |  | Set as 0 if none; 1 if any of 1-10 is true:  0.No 1.Yes | If 1 then col74 =1;  Set NAs as 0 |
| 53 | 162 | Ao.Arch.Procedure | 1.IPG+/-extension into the arch  2.IPG+reimplantation  3.Tube graft + sep. AVR  4.Bentall  5.VSRR  6.Homograft  7.Autograft/Ross  8. Aortic patch graft  9.SoV repair  10.Red aortoplasty  12.Extra-anatomic bypass |  | Set as 0 if none; 1 if any of 1-12 is true:  0.No 1.Yes | If 1 then col74 =1;  Set NAs as 0 |
| 54 | 163 | Ao.Desc.Procedure | 1.IPG+/-extension into the arch  2.IPG+reimplantation  3.Tube graft + sep. AVR  4.Bentall  5.VSRR  6.Homograft  7.Autograft/Ross  8. Aortic patch graft  9.SoV repair  10.Red aortoplasty  11. Concomitant stent  12.Extra-anatomic bypass |  | Set as 0 if none; 1 if any of 1-12 is true:  0.No 1.Yes | If 1 then col74 =1;  Set NAs as 0 |
| 55 | 164 | Ao.Abdo.Procedure | 1.IPG+/-extension into the arch  2.IPG+reimplantation  3.Tube graft + sep. AVR  4.Bentall  5.VSRR  6.Homograft  7.Autograft/Ross  8. Aortic patch graft  9.SoV repair  10.Red aortoplasty  11. Concomitant stent  12.Extra-anatomic bypass |  | mechanicalSupport |  |
| 56 | 175 | - mechanicalSupport   - Impeller   - Other.Mech.Support   - VAD   - IABP | 0.No  1.Preop  2.intraop  3.Post-op |  | Combine only Pre-op values 1. From all four variables into new variable, mechanical support:  0.No 1.Yes | Median.Sternotomy |
| 57 | 224 | Median.Sternotomy | 0.No 1.Yes |  |  | imdDecile |
| 58 | 225 | Partial.Sternotomy | 0.No 1.Yes |  |  | Set NAs to 0 |
| 59 | 226 | Thoracotomy | 0.No 1.Yes |  |  | Set NAs to 0 |
| 60 | 227 | Mini.Thoracotomy | 0.No 1.Yes |  |  | Set NAs to 0 |
| 61 |  | imdDecile | 1-10 |  | Same: 1 is most deprived | Set NAs to 1 |
|  |  | Mortality | 0.Alive  1.Dead |  | Same | Hypertension |
|  |  | OpDate | - | - |  |  |
|  |  | Year |  |  |  |  |

**Treatment of Missing Data**

The detailed methodology of data pre-processing and handling of missing data has been outlined in a previous study.[5] The overall percentage of missing data for baseline information is very low (1.7%). All patients with missing outcome values were excluded from the study so that any subsequent imputations on predictors do not modify the response variable. Age, Height and Weight were imputed using the median. For any other variables, the missing values were imputed by setting these to the baseline level of each variable, i.e., when no risks were present. For any variables with missing rate greater than 60%, such as LVF and Creatinine, we consulted with a experienced cardiac surgeon and confirmed that these clinical variable were not recorded (systematically missing) because these patient characteristics were considered to be normal, i.e. at baseline level by clinicians in the hospital. Hence, it was appropriate to set these variables using their baseline values.

Figure S1.1 CONSORT flow diagram for study.

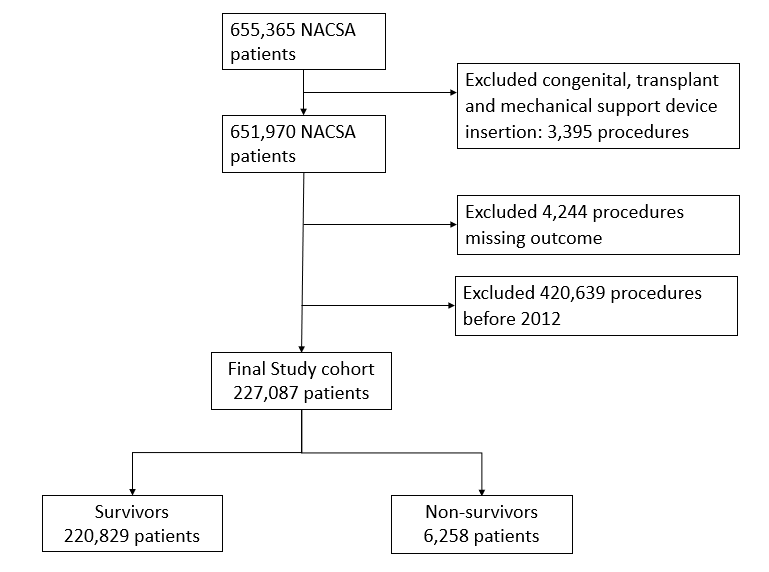

Figure S1.2 Variable missing rate for the variables considered. Some variables were derived from a combination of variables. Missing variables are backfilled to improve quality using other informative variables according to NACSA dataset cleaning protocol: <https://www.nicor.org.uk/wp-content/uploads/2018/09/nacsacleaning10.3.pdf>.

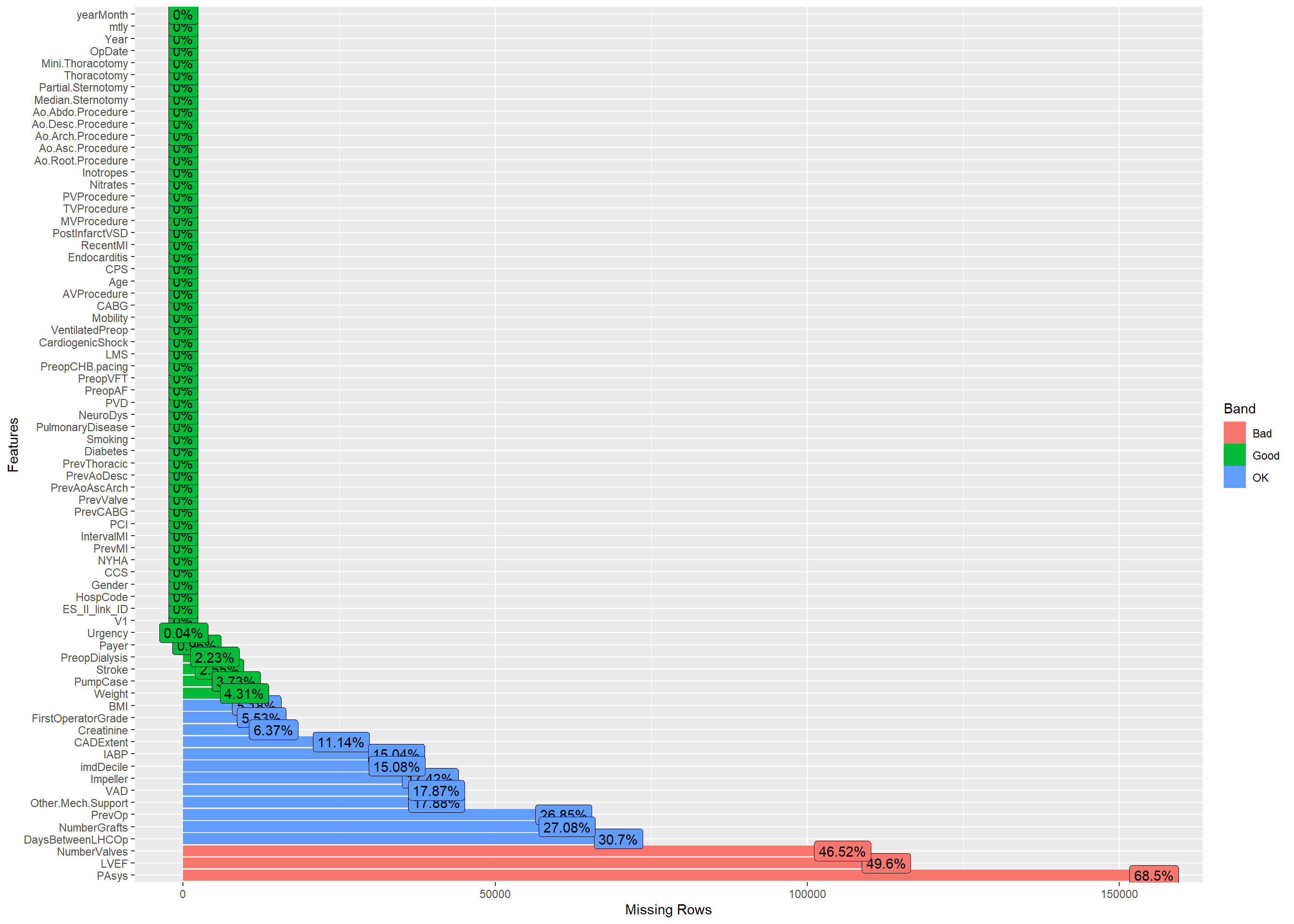

Figure S2. Flow diagram of baseline variable selection

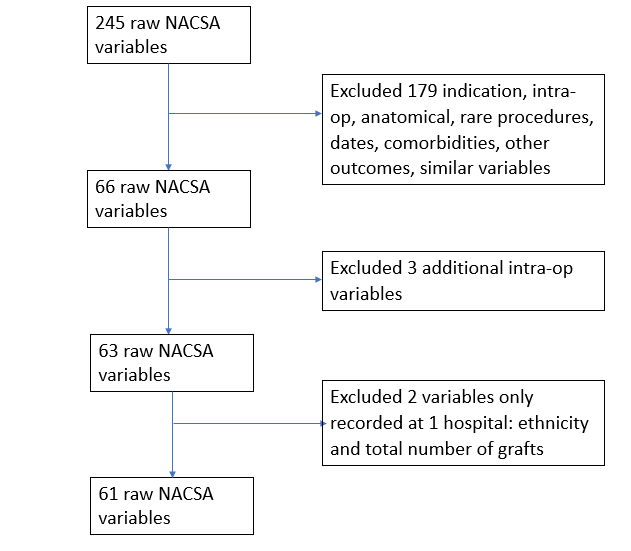

**Assessment of model performance**

The Area Under the Curve (AUC) performances of all variant models were evaluated, and the ROC curves plotted.[6] As a sensitivity analysis, we excluded the True Negative Rate from the performance evaluation, by calculating the F*_1_* score.[7] Decision Curve net benefit index is used to test clinical benefit.[8] 1 - Expected Calibration Error (ECE) was used to determine calibration performance, with higher values being better.[9] The adjusted Brier score (1 – Brier) was used without the normalization term,[10] but with higher values indicating better discrimination and calibration performance.

To determine the best model in terms of both discrimination and calibration, we took the geometric average of AUC, F1,[7] Decision Curve net benefit (Treated + Untreated), 1 – ECE and 1 – Brier. Geometric average has previously been found to be effective for summarising metrics for temporal based model calibration[11]. This metric is robust to outliers,[12] and is perferrable for aggregation compared to the weighted geometric mean.[13] The arithmetic average was used for Decision Curve net benefit over all thresholds as a measure of overall net benefit, before geometric averaging, since values can be negative.

Figure S3. Correlation analysis of all 61 variables

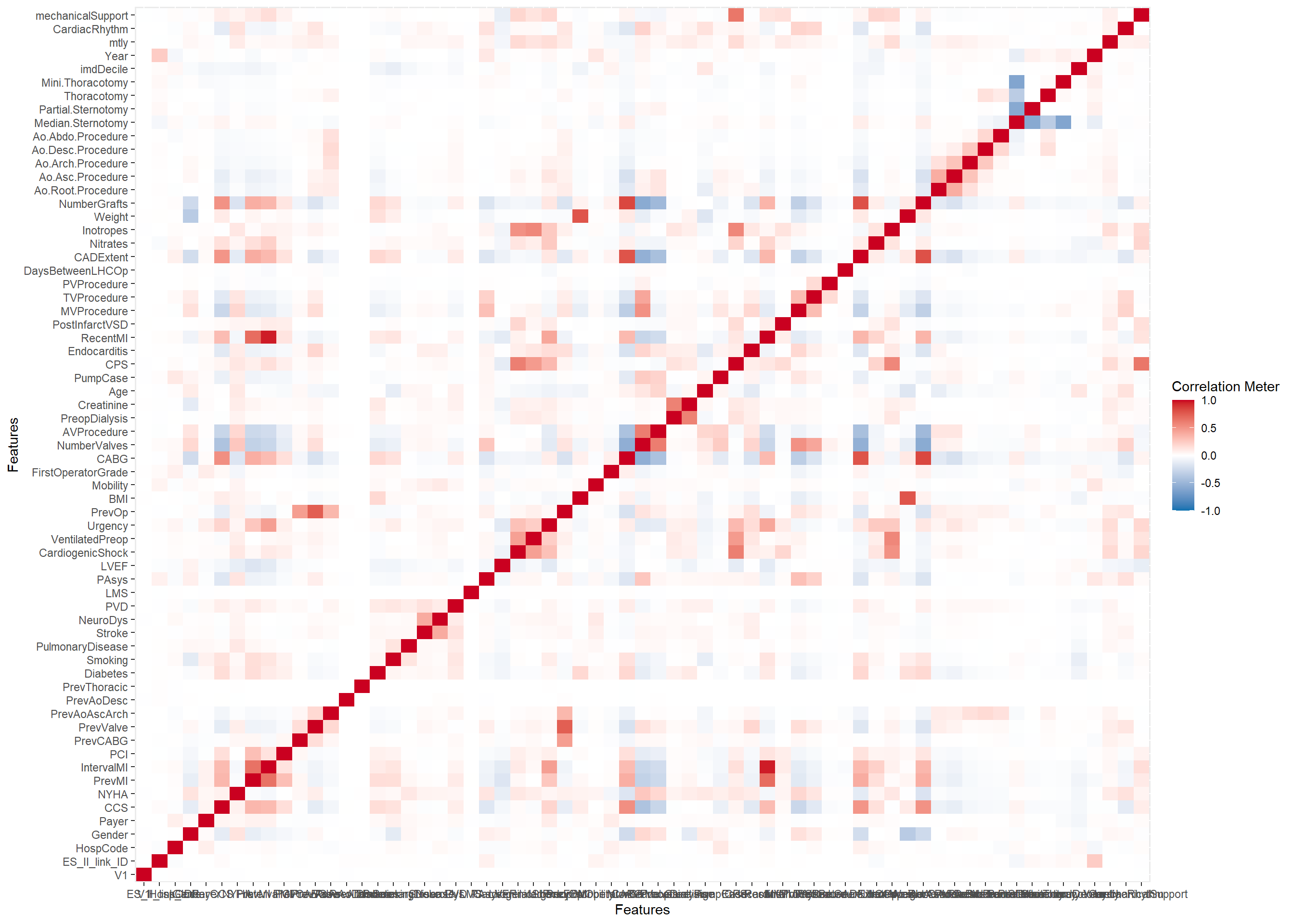

Figure S4. Data missing rate of variables after pre-processing.

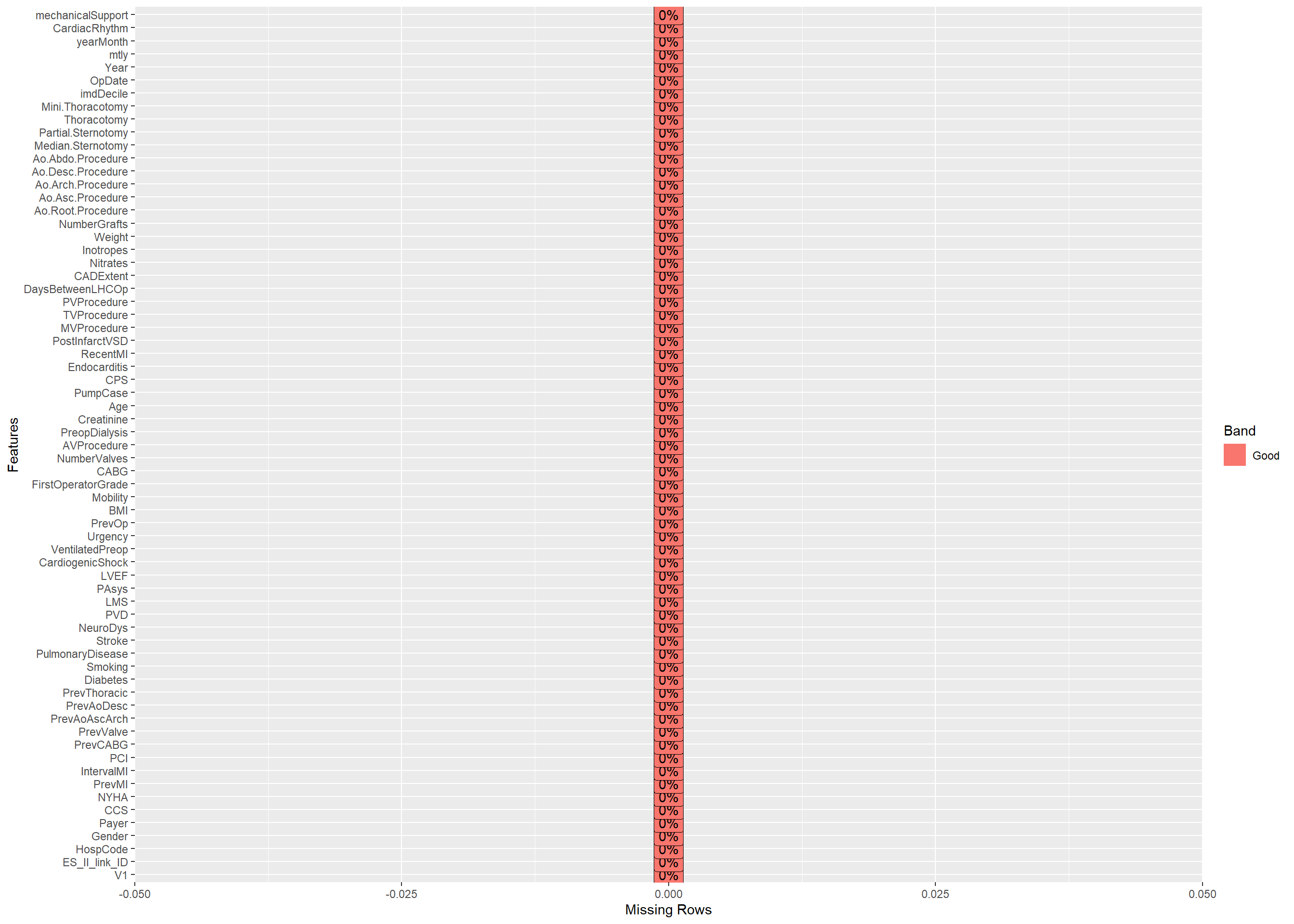

Table S2. Non-variable selected models: optimal model hyperparameters obtained from randomized 3-fold cross validation (CV) without variable selection. Mixed Effects Random Forest (MERF) and Mixed effects Xgboost models used the same initial hyperparameters as RF - Centre and Xgboost - Centre, respectively; - exclude variable, + include variable; RE refers to Random Effects.

| RF - Centre | n_estimators | 800 |
| --- | --- | --- |
|  | min_samples_split | 5 |
|  | min_samples_leaf | 10 |
|  | max_depth | 40 |
|  | bootstrap | TRUE |
| RF + Centre (hot-encoded) | n_estimators | 800 |
|  | min_samples_split | 12 |
|  | min_samples_leaf | 10 |
|  | max_depth | 60 |
|  | bootstrap | TRUE |
| Xgboost - Centre | subsample | 0.6 |
|  | n_estimators | 500 |
|  | min_child_weight | 5 |
|  | max_depth | 7 |
|  | gamma | 1 |
|  | colsample_bytree | 0.8 |
| Xgboost + Centre (hot-encoded) | subsample | 0.8 |
|  | n_estimators | 700 |
|  | min_child_weight | 0.5 |
|  | max_depth | 7 |
|  | gamma | 1.5 |
|  | colsample_bytree | 0.8 |
| GPBoost + RE: Centre | learning_rate | 0.5 |
|  | min_data_in_leaf | 50 |
|  | max_depth | 3 |
|  | max_bin | 2000 |
| Mixed Effects Xgboost + RE: centre | Xgboost – Centre values | See above |
|  | max_iterations | 5 |
| Mixed effects RF + RE: centre | RF – Centre values | See above |
|  | max_iterations | 5 |

Table S3. 18 Variable selected models: optimal model hyperparameters obtained from randomized 3-fold cross validation (CV) without variable selection. Mixed Effects Random Forest (MERF) and Mixed effects Xgboost models used the same initial hyperparameters as RF - Centre and Xgboost - Centre, respectively; - exclude variable, + include variable; RE refers to Random Effects.

| RF - Centre | n_estimators | 600 |
| --- | --- | --- |
|  | min_samples_split | 5 |
|  | min_samples_leaf | 40 |
|  | max_depth | 20 |
|  | bootstrap | True |
| RF + Centre (hot-encoded) | n_estimators | 600 |
|  | min_samples_split | 10 |
|  | min_samples_leaf | 20 |
|  | max_depth | 20 |
|  | bootstrap | True |
| Xgboost - Centre | subsample | 0.6 |
|  | n_estimators | 500 |
|  | min_child_weight | 5 |
|  | max_depth | 7 |
|  | gamma | 1 |
|  | colsample_bytree | 0.8 |
| Xgboost + Centre (hot-encoded) | subsample | 0.6 |
|  | n_estimators | 700 |
|  | min_child_weight | 1 |
|  | max_depth | 5 |
|  | gamma | 1.5 |
|  | colsample_bytree | 0.6 |
| GPBoost + Centre | learning_rate | 0.05 |
|  | min_data_in_leaf | 200 |
|  | max_depth | 5 |
|  | max_bin | 2000 |
|  | num_boost_round | 15 |
| Mixed Effects Xgboost (RE: centre) | subsample | 0.6 |
|  | n_estimators | 700 |
|  | min_child_weight | 1 |
|  | max_depth | 5 |
|  | gamma | 1.5 |
|  | colsample_bytree | 0.6 |
|  | max_iterations | 5 |
| Mixed effects RF (RE: centre) | n_estimators | 900 |
|  | min_samples_split | 12 |
|  | min_samples_leaf | 40 |
|  | max_depth | 20 |
|  | bootstrap | True |
|  | max_iterations | 5 |

Table S4 Mixed Effects Xgboost: top 18 important variables identified by Tree SHAP using Training dataset (n = 157196; 2012-2016);

| Rank | Variable id | Feature importance | Importance values |
| --- | --- | --- | --- |
| 1 | 23 | Urgency | 0.008594 |
| 2 | 33 | Age | 0.005628 |
| 3 | 32 | Creatinine | 0.005381 |
| 4 | 3 | NYHA | 0.003589 |
| 5 | 24 | PrevOp | 0.003184 |
| 6 | 19 | PAsys | 0.00186 |
| 7 | 35 | CPS | 0.001787 |
| 8 | 17 | PVD | 0.001655 |
| 9 | 58 | CardiacRhythm | 0.001446 |
| 10 | 21 | CardiogenicShock | 0.001282 |
| 11 | 39 | MVProcedure | 0.001136 |
| 12 | 0 | Gender | 0.001004 |
| 13 | 29 | NumberValves | 0.000996 |
| 14 | 46 | Weight | 0.000923 |
| 15 | 49 | Ao.Asc.Procedure | 0.000915 |
| 16 | 14 | PulmonaryDisease | 0.000741 |
| 17 | 31 | PreopDialysis | 0.000448 |
| 18 | 45 | Inotropes | 0.000428 |

Figure S5 Mixed Effects Xgboost: Tree SHAP feature importance plot for Training dataset (n = 157196; 2012-2016); every procedure is represented as a dot; the x position of the dot is the impact of that feature on the model’s prediction for that procedure in log-odds; procedures that do not fit on the row pile up to show regions of high case volume.

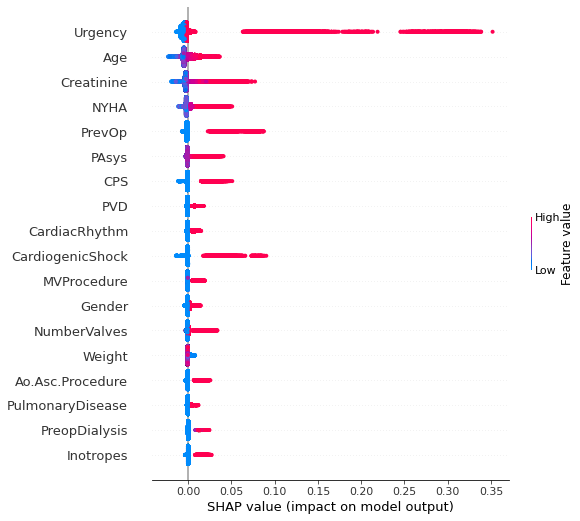

Figure S6 Mixed Effects Xgboost: mean absolute magnitude of importance across all prediction outputs;

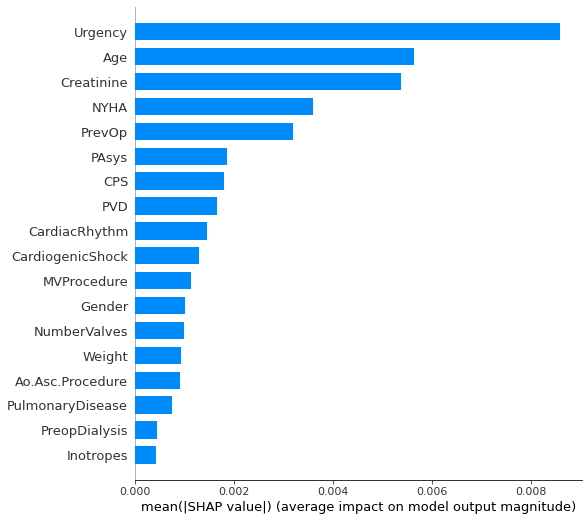

Table S5 Mixed Effects Random Forest (MERF): top 18 important variables identified by Tree SHAP using Training dataset (n = 157196; 2012-2016);

| Rank | Variable id | Feature importance | Importance values |
| --- | --- | --- | --- |
| 1 | 23 | Urgency | 0.011131 |
| 2 | 32 | Creatinine | 0.007791 |
| 3 | 33 | Age | 0.00734 |
| 4 | 3 | NYHA | 0.005258 |
| 5 | 24 | PrevOp | 0.004948 |
| 6 | 19 | PAsys | 0.002917 |
| 7 | 46 | Weight | 0.002785 |
| 8 | 35 | CPS | 0.002596 |
| 9 | 21 | CardiogenicShock | 0.002455 |
| 10 | 29 | NumberValves | 0.001961 |
| 11 | 17 | PVD | 0.001805 |
| 12 | 58 | CardiacRhythm | 0.001501 |
| 13 | 39 | MVProcedure | 0.001428 |
| 14 | 25 | BMI | 0.00139 |
| 15 | 47 | NumberGrafts | 0.001339 |
| 16 | 20 | LVEF | 0.000837 |
| 17 | 0 | Gender | 0.000757 |
| 18 | 42 | DaysBetweenLHCOp | 0.000735 |

Figure S7 Mixed Effects Random Forest (MERF): Tree SHAP feature importance plot for Training dataset (n = 157196; 2012-2016); every procedure is represented as a dot; the x position of the dot is the impact of that feature on the model’s prediction for that procedure in log-odds; procedures that do not fit on the row pile up to show regions of high case volume.

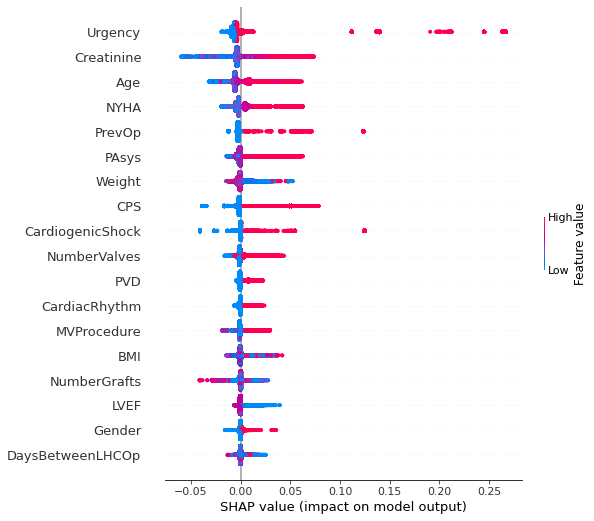

Figure S8 Mixed Effects Random Forest (MERF): mean absolute magnitude of importance across all prediction outputs;

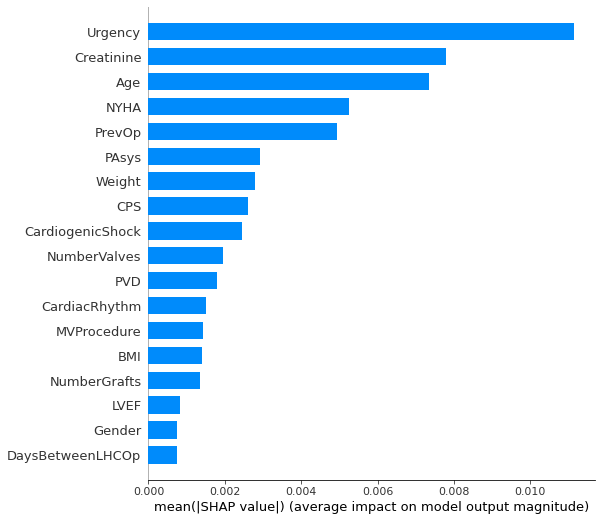

Table S6 Xgboost – Centre: top 18 important variables identified by Tree SHAP using Training dataset (n = 157196; 2012-2016);

| Rank | Variable id | Feature importance | Importance values |
| --- | --- | --- | --- |
| 1 | 33 | Age | 0.359045 |
| 2 | 23 | Urgency | 0.269227 |
| 3 | 32 | Creatinine | 0.233877 |
| 4 | 3 | NYHA | 0.225934 |
| 5 | 46 | Weight | 0.14223 |
| 6 | 17 | PVD | 0.09088 |
| 7 | 27 | FirstOperatorGrade | 0.090656 |
| 8 | 25 | BMI | 0.08896 |
| 9 | 58 | CardiacRhythm | 0.08348 |
| 10 | 24 | PrevOp | 0.083112 |
| 11 | 19 | PAsys | 0.077184 |
| 12 | 29 | NumberValves | 0.06959 |
| 13 | 14 | PulmonaryDisease | 0.064157 |
| 14 | 39 | MVProcedure | 0.062327 |
| 15 | 47 | NumberGrafts | 0.058565 |
| 16 | 35 | CPS | 0.050003 |
| 17 | 4 | PrevMI | 0.049979 |
| 18 | 0 | Gender | 0.047812 |

Figure S9 Xgboost - Centre: Tree SHAP feature importance plot for Training dataset (n = 157196; 2012-2016); every procedure is represented as a dot; the x position of the dot is the impact of that feature on the model’s prediction for that procedure in log-odds; procedures that do not fit on the row pile up to show regions of high case volume.

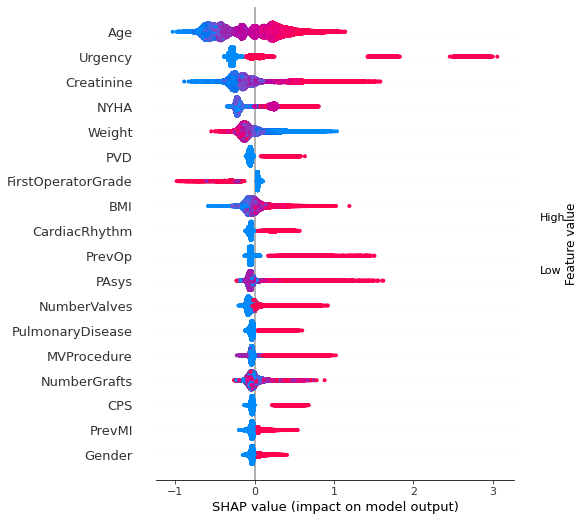

Figure S10 Xgboost - Centre: mean absolute magnitude of importance across all prediction outputs;

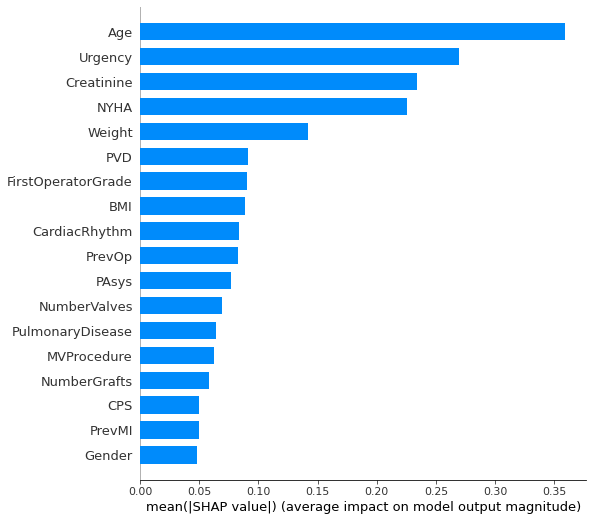

Table S7 Random Forest (RF) – Centre: top 18 important variables identified by Tree SHAP using Training dataset (n = 157196; 2012-2016);

| Rank | Variable id | Feature importance | Importance values |
| --- | --- | --- | --- |
| 1 | 23 | Urgency | 6.46E-03 |
| 2 | 32 | Creatinine | 6.31E-03 |
| 3 | 33 | Age | 6.16E-03 |
| 4 | 3 | NYHA | 4.61E-03 |
| 5 | 35 | CPS | 2.97E-03 |
| 6 | 19 | PAsys | 2.73E-03 |
| 7 | 46 | Weight | 2.72E-03 |
| 8 | 24 | PrevOp | 2.70E-03 |
| 9 | 58 | CardiacRhythm | 2.61E-03 |
| 10 | 17 | PVD | 2.16E-03 |
| 11 | 21 | CardiogenicShock | 2.13E-03 |
| 12 | 25 | BMI | 1.94E-03 |
| 13 | 39 | MVProcedure | 1.78E-03 |
| 14 | 29 | NumberValves | 1.65E-03 |
| 15 | 42 | DaysBetweenLHCOp | 1.47E-03 |
| 16 | 49 | Ao.Asc.Procedure | 1.44E-03 |
| 17 | 0 | Gender | 1.42E-03 |
| 18 | 45 | Inotropes | 1.40E-03 |

Figure S11 Random Forest (RF) - Centre: Tree SHAP feature importance plot for Training dataset (n = 157196; 2012-2016); every procedure is represented as a dot; the x position of the dot is the impact of that feature on the model’s prediction for that procedure in log-odds; procedures that do not fit on the row pile up to show regions of high case volume.

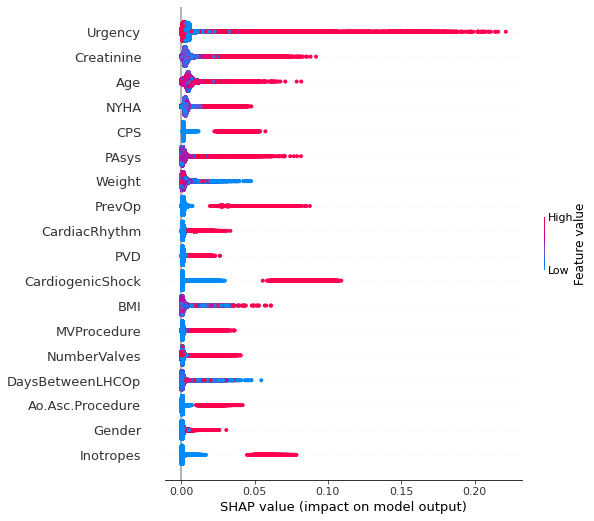

Figure S12 Random Forest (RF) - Centre: mean absolute magnitude of importance across all prediction outputs;

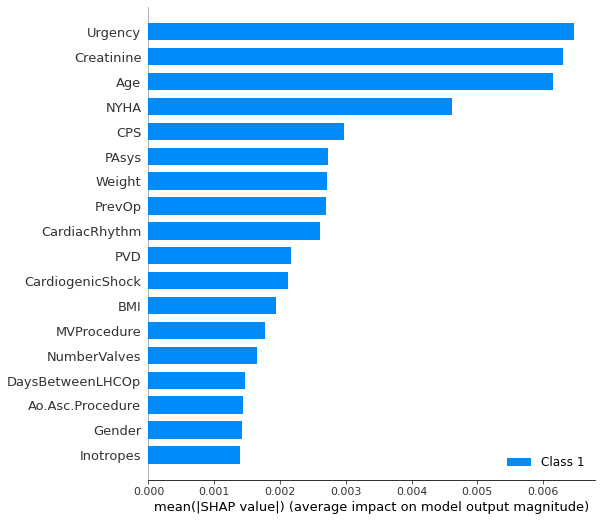

Table S8 Random Forest + Centre (hot-encoded): top 18 important variables identified by Tree SHAP using Training dataset (n = 157196; 2012-2016);

| Rank | Variable id | Feature importance | Importance values |
| --- | --- | --- | --- |
| 1 | 23 | Urgency | 0.006001113 |
| 2 | 32 | Creatinine | 0.005992479 |
| 3 | 33 | Age | 0.005627435 |
| 4 | 3 | NYHA | 0.004340263 |
| 5 | 35 | CPS | 0.002920918 |
| 6 | 19 | PAsys | 0.002622095 |
| 7 | 46 | Weight | 0.002563641 |
| 8 | 58 | CardiacRhythm | 0.002469642 |
| 9 | 24 | PrevOp | 0.002453433 |
| 10 | 17 | PVD | 0.002133912 |
| 11 | 21 | CardiogenicShock | 0.002056493 |
| 12 | 25 | BMI | 0.001735098 |
| 13 | 29 | NumberValves | 0.0016913 |
| 14 | 39 | MVProcedure | 0.001642142 |
| 15 | 49 | Ao.Asc.Procedure | 0.001516135 |
| 16 | 0 | Gender | 0.001389249 |
| 17 | 8 | PrevValve | 0.001349024 |
| 18 | 42 | DaysBetweenLHCOp | 0.001331966 |

Figure S13 Random Forest + Centre (hot-encoded): Tree SHAP feature importance plot for Training dataset (n = 157196; 2012-2016); every procedure is represented as a dot; the x position of the dot is the impact of that feature on the model’s prediction for that procedure in log-odds; procedures that do not fit on the row pile up to show regions of high case volume.

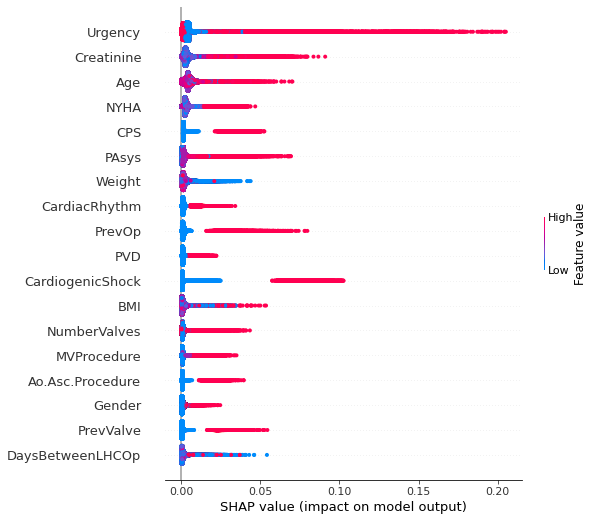

Figure S14 Random Forest + Centre (hot-encoded): mean absolute magnitude of importance across all prediction outputs;

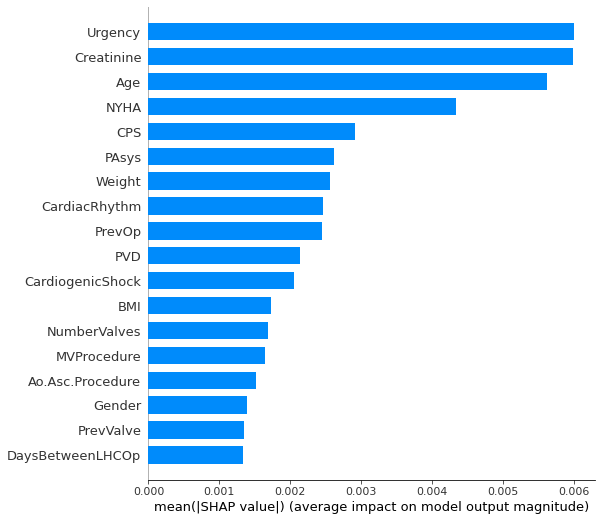

Table S9 Xgboost + Centre (hot-encoded): top 18 important variables identified by Tree SHAP using Training dataset (n = 157196; 2012-2016);

| Rank | Variable id | Feature importance | Importance values |
| --- | --- | --- | --- |
| 1 | 33 | Age | 0.34943044 |
| 2 | 23 | Urgency | 0.25036243 |
| 3 | 32 | Creatinine | 0.21957642 |
| 4 | 3 | NYHA | 0.19611274 |
| 5 | 46 | Weight | 0.1493066 |
| 6 | 25 | BMI | 0.089216046 |
| 7 | 27 | FirstOperatorGrade | 0.087769434 |
| 8 | 17 | PVD | 0.084330335 |
| 9 | 24 | PrevOp | 0.08148289 |
| 10 | 58 | CardiacRhythm | 0.08069693 |
| 11 | 29 | NumberValves | 0.06926664 |
| 12 | 14 | PulmonaryDisease | 0.06655002 |
| 13 | 39 | MVProcedure | 0.063090265 |
| 14 | 19 | PAsys | 0.062216528 |
| 15 | 0 | Gender | 0.061401874 |
| 16 | 4 | PrevMI | 0.056220867 |
| 17 | 35 | CPS | 0.050530296 |
| 18 | 49 | Ao.Asc.Procedure | 0.049323145 |

Figure S15 Xgboost + Centre (hot-encoded): Tree SHAP feature importance plot for Training dataset (n = 157196; 2012-2016); every procedure is represented as a dot; the x position of the dot is the impact of that feature on the model’s prediction for that procedure in log-odds; procedures that do not fit on the row pile up to show regions of high case volume.

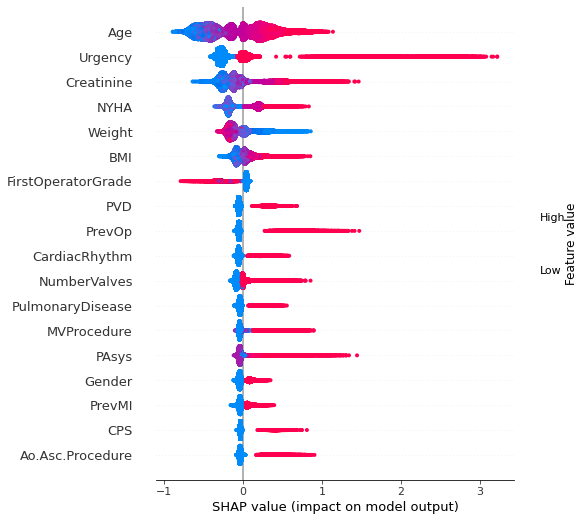

Figure S16 Xgboost + Centre (hot-encoded): mean absolute magnitude of importance across all prediction outputs;

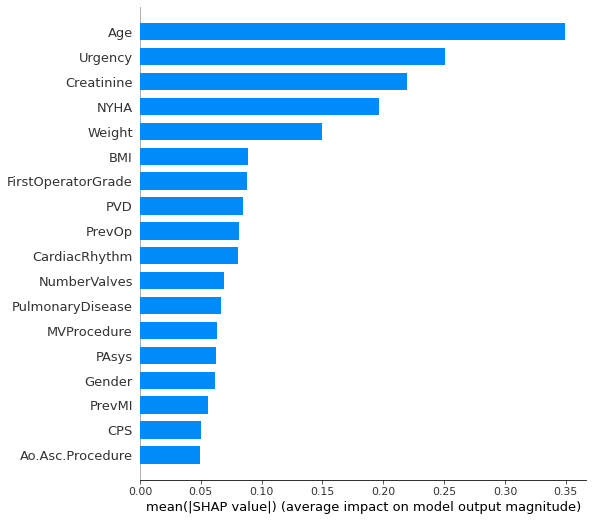

Table S10 GPBoost + Centre: top 18 important variables identified by Tree SHAP using Training dataset (n = 157196; 2012-2016);

| Rank | Variable id | Feature importance | Importance values |
| --- | --- | --- | --- |
| 1 | 23 | Urgency | 0.059172 |
| 2 | 33 | Age | 0.05241 |
| 3 | 32 | Creatinine | 0.045272 |
| 4 | 3 | NYHA | 0.042993 |
| 5 | 24 | PrevOp | 0.02459 |
| 6 | 35 | CPS | 0.013087 |
| 7 | 19 | PAsys | 0.009857 |
| 8 | 39 | MVProcedure | 0.008394 |
| 9 | 58 | CardiacRhythm | 0.006291 |
| 10 | 17 | PVD | 0.004586 |
| 11 | 21 | CardiogenicShock | 0.004415 |
| 12 | 29 | NumberValves | 0.004408 |
| 13 | 47 | NumberGrafts | 0.001617 |
| 14 | 31 | PreopDialysis | 0.001524 |
| 15 | 46 | Weight | 0.00148 |
| 16 | 50 | Ao.Arch.Procedure | 0.001167 |
| 17 | 4 | PrevMI | 0.001148 |
| 18 | 48 | Ao.Root.Procedure | 0.000977 |

Figure S17 GPBoost + Centre: Tree SHAP feature importance plot for Training dataset (n = 157196; 2012-2016); every procedure is represented as a dot; the x position of the dot is the impact of that feature on the model’s prediction for that procedure in log-odds; procedures that do not fit on the row pile up to show regions of high case volume.

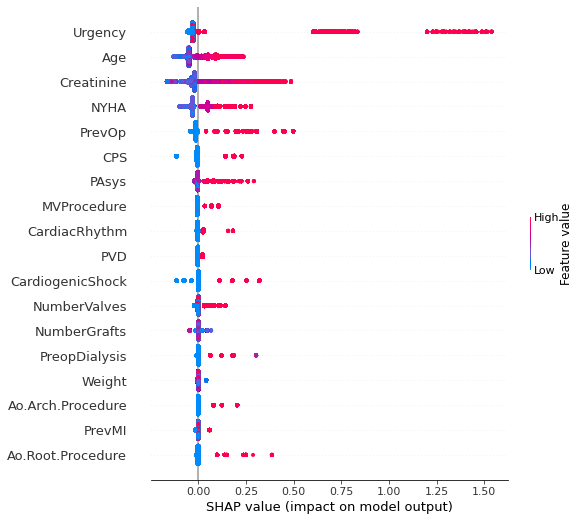

Figure S18 GPBoost + Centre: mean absolute magnitude of importance across all prediction outputs;

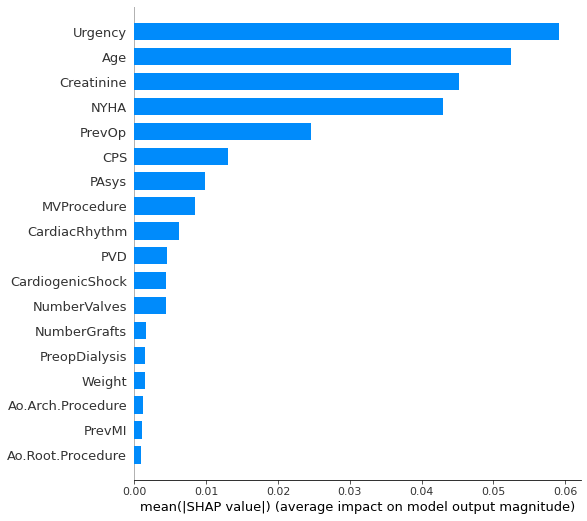

Table S11. Anova Test for non-variable selected models based on the CEM metric; ges - generalized eta squared is a measure of effect size

Effect	DFn	DFd	F p p<.05	ges

1	Model	6 6993 692402.4 0	* 0.998

Figure S19 Non-variable selected model performances are compared using multiple pairwise paired t-tests with Bonferroni correction.

Table S12. Top 18 important variables for each of the non-variable selected models; ordered by importance ranking.

| Rank | Xgboost – Centre | Xgboost + Centre (hot-encoded) | Mixed Effects Xgboost | Random Forest (RF) – Centre | Random Forest + Centre (hot-encoded) | Mixed Effects Random Forest (MERF) | GPBoost + Centre |
| --- | --- | --- | --- | --- | --- | --- | --- |
| 1 | Age | Age | Urgency | Urgency | Urgency | Urgency | Urgency |
| 2 | Urgency | Urgency | Age | Creatinine | Creatinine | Creatinine | Age |
| 3 | Creatinine | Creatinine | Creatinine | Age | Age | Age | Creatinine |
| 4 | NYHA | NYHA | NYHA | NYHA | NYHA | NYHA | NYHA |
| 5 | Weight | Weight | PrevOp | CPS | CPS | PrevOp | PrevOp |
| 6 | PVD | BMI | PAsys | PAsys | PAsys | PAsys | CPS |
| 7 | FirstOperatorGrade | FirstOperatorGrade | CPS | Weight | Weight | Weight | PAsys |
| 8 | BMI | PVD | PVD | PrevOp | CardiacRhythm | CPS | MVProcedure |
| 9 | CardiacRhythm | PrevOp | CardiacRhythm | CardiacRhythm | PrevOp | CardiogenicShock | CardiacRhythm |
| 10 | PrevOp | CardiacRhythm | CardiogenicShock | PVD | PVD | NumberValves | PVD |
| 11 | PAsys | NumberValves | MVProcedure | CardiogenicShock | CardiogenicShock | PVD | CardiogenicShock |
| 12 | NumberValves | PulmonaryDisease | Gender | BMI | BMI | CardiacRhythm | NumberValves |
| 13 | PulmonaryDisease | MVProcedure | NumberValves | MVProcedure | NumberValves | MVProcedure | NumberGrafts |
| 14 | MVProcedure | PAsys | Weight | NumberValves | MVProcedure | BMI | PreopDialysis |
| 15 | NumberGrafts | Gender | Ao.Asc.Procedure | DaysBetweenLHCOp | Ao.Asc.Procedure | NumberGrafts | Weight |
| 16 | CPS | PrevMI | PulmonaryDisease | Ao.Asc.Procedure | Gender | LVEF | Ao.Arch.Procedure |
| 17 | PrevMI | CPS | PreopDialysis | Gender | PrevValve | Gender | PrevMI |
| 18 | Gender | Ao.Asc.Procedure | Inotropes | Inotropes | DaysBetweenLHCOp | DaysBetweenLHCOp | Ao.Root.Procedure |

Table S13. Anova Test for variable selected models based on the CEM metric; ges - generalized eta squared is a measure of effect size

Effect	DFn	DFd	F p p<.05	ges

1	Model	6 6993 553210.7 0	* 0.998

Figure S20 Variable selected model performances are compared using multiple pairwise paired t-tests with Bonferroni correction.

Table S14. Variable selected models: Dunnett's test with Xgboost - Centre as control; 95% family-wise confidence level are shown as well as mean difference in CEM and p-values; NC: no centre; HE: hot-encoded centre; ME: mixed effects.

|  |  |  |  | **95% CI** | |
| --- | --- | --- | --- | --- | --- |
| **Group 1** | **Group 2** | **CEM Difference (1-2)** | ***P* Value** | **Lower Bound** | **Upper Bound** |
| GPBoost ME | Xgboost NC (Control) | -0.0667 | <2e-16 | -0.0672 | -0.0661 |
| RF HE |  | -0.0071 | <2e-16 | -0.0077 | -0.0065 |
| RF ME |  | -0.2535 | <2e-16 | -0.2540 | -0.2529 |
| RF NC |  | -0.0075 | <2e-16 | -0.0081 | -0.0070 |
| Xgboost HE |  | 0.0002 | 0.9437 | -0.0004 | 0.0007 |
| Xgboost ME |  | -0.2539 | <2e-16 | -0.2545 | -0.2533 |

Signif. codes: 0 '***' 0.001 '**' 0.01 '*' 0.05 '.' 0.1 ' '

Table S15. Variable selected models: Geometric Mean of Individual metrics; CEM refs to Clinical Effective Metric; Standard deviation and 95% CI are shown for CEM; adjusted 1 - ECE and 1 - Brier score values are shown; net benefit is average absolute overall benefit across all thresholds.

| **Model Category** | **ECE** | **AUC** | **Brier** | **F1** | **Net Benefit** | **CEM Mean** | **CEM S.D** | **CEM Lower CI** | **CEM Upper CI** |
| --- | --- | --- | --- | --- | --- | --- | --- | --- | --- |
| GPBoost ME | 0.989 | 0.719 | 0.974 | 0.209 | 0.887 | 0.664 | 0.007 | 0.663 | 0.664 |
| RF HE | 0.998 | 0.832 | 0.976 | 0.270 | 0.904 | 0.723 | 0.005 | 0.723 | 0.724 |
| RF ME | 0.520 | 0.826 | 0.744 | 0.266 | 0.290 | 0.477 | 0.004 | 0.477 | 0.477 |
| RF NC | 0.997 | 0.832 | 0.976 | 0.270 | 0.904 | 0.723 | 0.005 | 0.722 | 0.723 |
| Xgboost HE | 0.998 | 0.841 | 0.976 | 0.280 | 0.905 | 0.730 | 0.005 | 0.730 | 0.731 |
| Xgboost ME | 0.520 | 0.825 | 0.744 | 0.264 | 0.291 | 0.476 | 0.004 | 0.476 | 0.477 |
| Xgboost NC | 0.998 | 0.840 | 0.977 | 0.280 | 0.906 | 0.730 | 0.005 | 0.730 | 0.731 |

Table S16. Anova Test for Xgboost (ES II) against the best non-variable and variable selected models based on the CEM metric; ges - generalized eta squared is a measure of effect size

Effect	DFn	DFd	F p p<.05	ges

1	Model	2 2997 2427.263 0	* 0.618

Figure S21 Variable selected models: adapted Rain plot of CEM constituent metrics by model; larger sized spheres represent higher metric performance and vice versa. ECE: 1-ECE; Brier: 1-Brier.

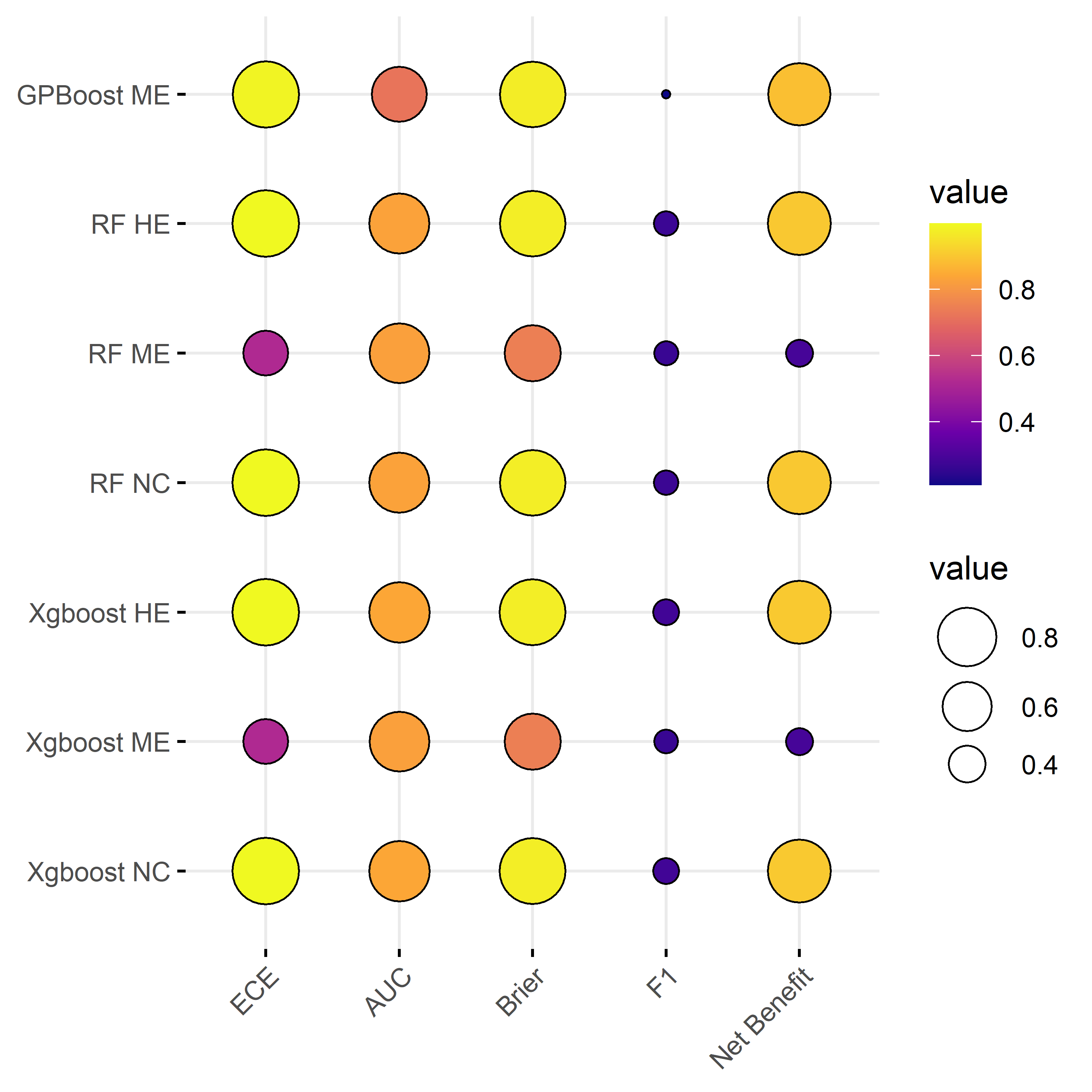

Table S17. Dunnett's test comparing: Xgboost (ES II), the best non-variable and variable selected models, with Xgboost (ES II variables) model as control; 95% family-wise confidence level are shown as well as mean difference in CEM and p-values; HE: hot-encoded centre; NVS: non-variable selected; VS: variable selected; ES II: EuroSCORE II.

|  |  |  |  | **95% CI** | |
| --- | --- | --- | --- | --- | --- |
| **Group 1** | **Group 2** | **CEM Difference (1-2)** | ***P* Value** | **Lower Bound** | **Upper Bound** |
| Xgboost HE (NVS) | Xgboost (ES II) | 0.0150 | < 2e-16 *** | 0.0145 | 0.0155 |
| Xgboost HE (VS) |  | 0.0023 | 6.3e-15 *** | 0.0018 | 0.0028 |

Signif. codes: 0 '***' 0.001 '**' 0.01 '*' 0.05 '.' 0.1 ' '
